# Plasma Proteomic Identifies GDF15 as a Candidate Lithium Treatment Response Biomarker in Mild Cognitive Impairment: A Secondary Analysis of the LATTICE Randomized Clinical Trial

**DOI:** 10.64898/2026.11.14.26368038

**Authors:** Anuradha Sehrawat, Ruyu Shi, Marissa F. Farinas, James E. Emanuel, Xuemei Zeng, Brian J. Lopresti, Sarah K. Royse, Andrea M. Weinstein, Meryl A. Butters, Laisze Lee, Ariel G. Gildengers, Thomas K. Karikari

## Abstract

Lithium (Li) demonstrates neuroprotective effects relevant to Alzheimer’s disease (AD); however, its molecular mechanisms remain insufficiently characterized. This study analyzed plasma proteomic profiles from the Li as a Treatment to Prevent Impairment of Cognition in Elders (LATTICE) randomized clinical trial to identify markers associated with amyloid (Aβ) pathology, disease progression, and response to Li treatment.

In the LATTICE trial, 80 older adults with mild cognitive impairment (MCI) were randomly assigned to receive either low-dose Li carbonate or placebo for two years. Longitudinal plasma samples were analyzed using the NULISAseq CNS Disease Panel 120 proteomic panel. Baseline protein levels were compared according to randomized treatment assignment, Aβ-PET status, and MCI subtype. Analyses included longitudinal changes in protein levels associated with treatment and time, associations between baseline protein levels and subsequent cognitive, functional, and neuroimaging trajectories, and the influence of baseline protein levels on Li treatment effects.

Among the 80 participants, 62 had amnestic MCI and 21 were Aβ-PET positive. Baseline proteomic differences were more strongly associated with Aβ-PET status than with randomized treatment assignment. Plasma p-tau217 concentrations were elevated in Aβ-PET positive individuals and remained significant after false discovery rate correction. Over time, CALB2 and TREM2 exhibited nominal treatment-by-time interactions in the full cohort, while in the amnestic MCI subgroup, CALB2, TREM2, and Aβ40 demonstrated nominal effects; none persisted after multiple-testing correction. GDF15 showed the most consistent associations with CDR Sum of Boxes trajectories and treatment-effect modification that remained significant after false discovery rate correction in the amnestic MCI subgroup, whereas p-tau217 was most consistently associated with hippocampal volume trajectories. Other protein-outcome associations were exploratory.

Plasma proteomics identified distinct molecular patterns associated with Aβ pathology and the MCI phenotype, along with exploratory evidence of Li-associated protein changes. Plasma GDF15 and p-tau217 emerged as potential markers of clinical progression and treatment-response heterogeneity, warranting validation in larger, adequately powered trials.

## Introduction

Lithium (Li) maintains neural homeostasis at low physiological concentrations ^1,2^. In mild cognitive impairment (MCI) due to Alzheimer’s disease (AD), amyloid-β (Aβ) deposits sequester Li, resulting in region-specific depletion and reduced bioavailability ^1^. Disrupted Li homeostasis is considered an early and potentially modifiable contributor to AD ^1^. Preclinical and translational studies demonstrate that Li reduces Aβ production and promotes its clearance by inhibiting BACE1 and γ-secretase, as well as enhancing LRP1-mediated efflux ^3–5^. Li may also attenuate tau pathology by inhibiting GSK-3β, decreasing tau expression, and increasing tau ubiquitination ^6–8^. Other proposed neuroprotective mechanisms include the activation of Wnt/β-catenin signaling, autophagy, neurogenesis, and BDNF expression; preservation of mitochondrial and synaptic function; and reduction of oxidative stress and neuroinflammation ^9–12^.

Epidemiological studies have associated long-term therapeutic Li use and higher environmental Li exposure with lower dementia incidence ^13–16^. However, clinical evidence remains inconclusive. A 2024 meta-analysis reported reduced risks of all-cause dementia and AD among individuals receiving Li therapy and identified an association between longer treatment duration and lower dementia incidence ^17^. Longer-term trials of low-dose Li in amnestic MCI and early or mild AD dementia have demonstrated cognitive stabilization and changes in CSF biomarkers, including reduced phosphorylated tau and increased Aβ1-42 ^18,19^. A network meta-analysis of eight randomized placebo-controlled trials found that low-dose lithium improved MMSE scores^20^. A 2026 systematic review provided preliminary evidence that longer-term Li treatment may slow cognitive decline in MCI and early dementia, particularly in studies with extended treatment durations ^21^. In contrast, other randomized trials and meta-analyses have reported minimal or no cognitive benefit ^22–24^. A broader review of 30 studies with 727 participants also found varied cognitive effects across populations and study designs ^25^. Collectively, variability in Li exposure, treatment duration, disease stage, study population, and outcome measures underscore the need to better characterize Li-associated molecular responses in humans.

Recent advances in high-sensitivity proteomics enable simultaneous measurement of many central nervous system (CNS)-related protein biomarkers from small plasma samples ^26^. For instance, the NULISAseq CNS Disease Panel quantifies approximately 120 or more proteins associated with Aβ and tau pathology, neurodegeneration, inflammation, vascular dysfunction, and synaptic biology ^26,27^. This platform facilitates identification of molecular signatures linked to disease progression and potential treatment response. When applied to longitudinal therapeutic trial samples, it enables characterization of treatment-associated molecular changes and their relationship to clinical outcomes ^26–28^.

The Li as a Treatment to Prevent Impairment of Cognition in Elders (LATTICE) pilot randomized controlled trial evaluated whether two years of low-dose Li carbonate could slow cognitive decline and structural brain changes in older adults with MCI. The trial established the feasibility, safety, and tolerability of low-dose Li and provided preliminary evidence of a slower decline in verbal memory, thereby supporting further research into the clinical and biological effects of Li in MCI ^29,30^. Building on this trial, the present secondary analysis used the NULISAseq CNS panel to characterize baseline protein differences by treatment assignment, Aβ-PET status, and MCI subtype; assess protein trajectories during Li treatment; and examine associations between baseline proteins and subsequent cognitive, functional, and imaging outcomes. Additionally, we evaluated whether baseline proteins could identify Li treatment effects, with the objective of identifying candidate prognostic and treatment-response biomarkers in MCI.

## Materials and methods

### Study Design

LATTICE is a single-site, randomized, double-blind, placebo-controlled clinical trial at the University of Pittsburgh School of Medicine (IRB-approved; CONSORT-compliant). Enrollment began on September 1, 2017, supervised by an independent Data Safety and Monitoring Board (DSMB). The full trial design and primary outcomes have been described previously ^29,30^. Eligible participants were adults aged 60 or older with MCI, defined by Petersen criteria as performance 1-2 standard deviations below age- and education-adjusted norms in at least one cognitive domain. All participants had preserved activities of daily living and no major psychiatric or neurologic comorbidities. Enrollment took place between February 2018 and August 2024. Participants were randomized 1:1 to receive daily oral Li carbonate (150-300 mg, titrated weekly to maximum tolerated dose) or identical placebo, using permuted blocks stratified by Aβ-beta (Aβ) plaque status. Assessments occurred at baseline (T1), Year 1 (T2), and Year 2 (T3). These included a neuropsychological battery (adapted PACC, CVLT-II, BVMT-R), 7-Tesla structural MRI processed with FreeSurfer v8, simultaneous [¹¹C]-PiB PET/MRI to quantify cerebral Aβ burden (positivity threshold: GBL9 SUVR ≥1.346), and standardized clinical measures of comorbidity (CIRS-G), cardiovascular risk (Framingham Stroke Risk Profile), mood (PHQ-9), physical activity (PASE), adherence (BARS), and adverse effects (UKU Scale). APOE genotype was determined from baseline samples.

### NULISAseq CNS Disease Panel 120 Assay

Available longitudinal plasma samples were analyzed using the NULISAseq CNS Disease Panel 120 assay (Alamar Biosciences) according to the manufacturer’s instructions. Target proteins were captured using antibody pairs coupled to DNA reporters. Following immunocomplex formation and sequential capture, wash, and release steps, DNA reporters were ligated, pooled, amplified, purified, and sequenced. Sequencing data were processed using the NULISAseq workflow to generate normalized, log2-transformed NULISA Protein Quantification (NPQ) values.

### Statistical Analysis

All analyses were performed in R version 4.5.1 (R Foundation for Statistical Computing, Vienna, Austria). NULISA NPQ values were analyzed on the log2 scale, and replicate measurements from the same participant and visit were averaged on the log2 scale. Baseline protein analyses included participants with available baseline plasma (n = 73; 36 Li and 37 placebo), and longitudinal analyses included all available plasma samples from T1 to T3 (205 samples from 80 participants). Unless otherwise specified, tests were two-sided, and nominal significance was defined as an unadjusted p < 0.05. Multiple testing across the 127 proteins was controlled using the Benjamini-Hochberg false discovery rate (FDR), with FDR < 0.05 considered significant. Baseline characteristics were summarized as medians with interquartile ranges (IQRs) for continuous variables and as counts and percentages for categorical variables. Differences by MCI subtype, treatment assignment, Aβ-PET status, and treatment-by-PiB subgroup were assessed using the Kruskal-Wallis test for continuous variables and Pearson’s χ^2^ test or Fisher’s exact test for categorical variables as appropriate.

Baseline differences in protein levels by treatment assignment (Li versus placebo), Aβ-PET status (PiB+ versus PiB-), and MCI subtype (aMCI versus non-aMCI) were evaluated using linear models adjusted for age and sex, fitted with the limma package (version 3.64.3) with empirical Bayes moderation of variances (intensity trend and robust estimation). For the four treatment-by-PiB subgroups (placebo/PiB-, placebo/PiB+, Li/PiB-, and Li/PiB+), subgroup was modeled as a four-level factor, and all six pairwise contrasts were estimated from the same model. Regression coefficients are reported on the log_2_ scale and as fold changes (FC = 2^coefficient), with FDR correction applied across proteins within each contrast. For descriptive purposes, unadjusted differences were also assessed using the Wilcoxon rank-sum test (two groups) or the Kruskal-Wallis test (four subgroups), with fold changes calculated from group medians (FC = 2^[difference in median log2 NPQ]); these unadjusted statistics are annotated on the box plots. The ability of single proteins to discriminate treatment assignment (overall and within PiB+ and PiB-subgroups), aMCI from non-aMCI, and PiB status among participants with aMCI was assessed using receiver operating characteristic (ROC) analysis with the pROC package (version 1.19.0.1); 95% CIs for the area under the curve (AUC) were derived from 1,000 bootstrap resamples stratified by group.

Longitudinal changes in plasma proteins were evaluated using linear mixed-effects models (lme4 version 2.0-1 and lmerTest version 3.2-1) with protein NPQ as the outcome and treatment, study visit (coded 0, 1, and 2 for T1, T2, and T3), the treatment-by-time interaction, baseline age, and sex as fixed effects. Models included participant-specific random intercepts and random slopes for visit, with random slopes removed when that model was singular or failed to converge. The treatment-by-time interaction, estimated with restricted maximum likelihood and Satterthwaite-approximated degrees of freedom, tested whether protein trajectories differed between treatment groups; its coefficient (β) represents the difference between the Li and placebo groups in change in log2 NPQ per visit interval. Analyses were repeated in participants with aMCI at baseline. Associations between baseline protein levels and subsequent trajectories of eight outcomes (CDR Sum of Boxes, PASE, PACC, PHQ-9, BVMT-R Delayed Recall, CVLT-II Long-Delay Free Recall, hippocampal volume, and cortical volume) were assessed in participants with known Aβ-PET status using linear mixed-effects models with the same random-effects structure. Baseline protein levels were z-scored within each analytic sample, and all models were adjusted for baseline PiB status, age, and sex. Three models were fitted for each protein and outcome. The standalone prognostic model included the baseline protein-by-time interaction and tested whether baseline protein level was associated with the rate of change in the outcome (Wald test). The add-on prognostic model tested whether adding the baseline protein main effect and protein-by-time term to a model already containing the treatment-by-time interaction improved model fit (likelihood ratio test). The predictive model included the baseline protein-by-treatment-by-time interaction with all lower-order terms and tested whether baseline protein level modified the effect of Li on the outcome trajectory (likelihood ratio test comparing models with and without the three-way term). Models compared using likelihood ratio tests were fitted by maximum likelihood. To allow comparison across outcomes, coefficients were divided by the standard deviation of the outcome, yielding standardized β values expressed as the SD change in the outcome per visit interval per 1-SD higher baseline protein level (for predictive models, the difference in this association between the Li and placebo groups). All models were fitted in the full cohort and in the aMCI subgroup, and FDR correction was applied across proteins separately for each outcome, model, and population. For visualization, model-predicted outcome trajectories by treatment group were estimated from the predictive model at 1 SD below and above the mean baseline protein level using estimated marginal means (emmeans version 2.0.3), holding age at its mean and averaging over sex and PiB status in proportion to the analytic sample; Li-placebo differences in slope (per visit interval) at each protein level were estimated with Satterthwaite-approximated degrees of freedom.

## Results

### Baseline Characteristics of Study Participants

The study cohort comprised 80 participants, with 62 (77.5%) classified as having amnestic MCI (aMCI) and 18 (22.5%) as non-amnestic MCI (non-aMCI; Table 1). The median age was 71.5 years (interquartile range [IQR], 66.5-78.2), and the median duration of education was 16 years (IQR, 14-18). Females constituted 56.3% of the cohort, and 88.8% identified as non-Hispanic White. Participants with aMCI demonstrated lower median scores on the Preclinical Alzheimer Cognitive Composite (PACC; 0.3 versus 1.8; p = 0.009), the Brief Visuospatial Memory Test-Revised (BVMT-R) Delayed Recall (5.0 versus 8.0; p = 0.011), and the California Verbal Learning Test-II (CVLT-II) Long-Delay Free Recall (7.0 versus 12.0; p□=□8.79□×□10-□) compared to those with non-aMCI. Aβ-PET status was available for 75 participants; 21 were PiB-positive (28.0% of those with available PET status and 26.3% of the full cohort), and 5 had missing PET status. All 21 PiB-positive participants had aMCI. Aβ-PET status differed by MCI subtype (p = 0.003). We observed no nominal differences between MCI groups for other demographic, clinical, or brain volume measures.

**Table 1.** Demographic and clinical characteristics of study participants by MCI subtype

| **Characteristic** | **N** | **Overall**<br>**N = 80** | **Amnestic MCI**<br>**N = 62** | **Non-amnestic MCI**<br>**N = 18** | **p-value** |
|---|---|---|---|---|---|
| **Age**, years | 80 | 71.5 (66.5, 78.2) | 71.8 (66.9, 78.4) | 68.9 (63.1, 75.6) | 0.249 |
| **Education**, years | 80 | 16.0 (14.0, 18.0) | 16.0 (15.0, 18.0) | 16.0 (14.0, 17.0) | 0.282 |
| **Burden score** | 80 | 1.5 (0.0, 4.0) | 2.0 (0.0, 4.0) | 1.0 (1.0, 5.5) | 0.707 |
| **eGFR (CKD-EPI 2021)** | 80 | 84.2 (71.4, 91.9) | 85.6 (71.8, 92.4) | 77.3 (69.2, 85.8) | 0.148 |
| **Serum creatinine**, mg/dL | 80 | 0.9 (0.8, 1.0) | 0.9 (0.8, 1.0) | 0.9 (0.7, 1.0) | 0.729 |
| **Number of medications** | 80 | 6.0 (4.0, 9.0) | 6.0 (4.0, 9.0) | 6.0 (5.0, 9.0) | 0.677 |
| **CDR Sum of Boxes** | 80 | 0.5 (0.3, 1.0) | 0.8 (0.5, 1.5) | 0.5 (0.0, 1.0) | 0.477 |
| **PHQ-9 total score** | 80 | 3.0 (1.0, 5.0) | 3.0 (1.0, 5.0) | 3.0 (1.0, 8.0) | 0.767 |
| **PASE total score** | 79 | 92.1 (61.8, 133.3) | 92.3 (61.1, 127.2) | 83.2 (69.6, 149.6) | 0.512 |
| **PACC** | 77 | 0.8 (-2.3, 2.0) | 0.3 (-2.6, 1.4) | 1.8 (0.8, 4.0) | 0.009 |
| **Hippocampal volume**, mm<sup>3</sup> | 62 | 7,392.4 (6,630.7, 7,749.3) | 7,418.0 (6,630.7, 7,766.2) | 7,337.0 (6,615.9, 7,721.2) | 0.818 |
| **BVMT-R Delayed Recall** | 79 | 7.0 (4.0, 9.0) | 5.0 (4.0, 8.0) | 8.0 (7.0, 10.0) | 0.011 |
| **CVLT-II Long-Delay Free Recall** | 79 | 8.0 (6.0, 10.0) | 7.0 (5.0, 9.0) | 12.0 (9.0, 14.0) | 8.79 × 10<sup>-6</sup> |
| **Cortical volume**, mm<sup>3</sup> | 62 | 408,790.2 (397,107.8, 428,692.6) | 407,397.5 (397,107.8, 428,692.6) | 420,939.1 (392,357.7, 431,097.9) | 0.490 |
| **Sex** | 80 |  |  |  | 0.121 |
| Female |  | 45 (56.3%) | 32 (51.6%) | 13 (72.2%) |  |
| Male |  | 35 (43.8%) | 30 (48.4%) | 5 (27.8%) |  |
| **Race** | 80 |  |  |  | 0.283 |
| Asian |  | 1 (1.3%) | 0 (0.0%) | 1 (5.6%) |  |
| Black or African American |  | 8 (10.0%) | 6 (9.7%) | 2 (11.1%) |  |
| White |  | 71 (88.8%) | 56 (90.3%) | 15 (83.3%) |  |
| **Ethnicity** | 80 |  |  |  | 1.000 |
| Not Hispanic or Latino |  | 79 (98.8%) | 61 (98.4%) | 18 (100.0%) |  |
| Hispanic or Latino |  | 1 (1.3%) | 1 (1.6%) | 0 (0.0%) |  |
| **Aβ-PET status** | 80 |  |  |  | **0.003** |
| PiB- |  | 54 (67.5%) | 38 (61.3%) | 16 (88.9%) |  |
| PiB+ |  | 21 (26.3%) | 21 (33.9%) | 0 (0.0%) |  |
| Unknown |  | 5 (6.3%) | 3 (4.8%) | 2 (11.1%) |  |
| **CDR Global score** | 80 |  |  |  | 0.770 |
| 0 |  | 29 (36.3%) | 23 (37.1%) | 6 (33.3%) |  |
| 0.5 |  | 51 (63.8%) | 39 (62.9%) | 12 (66.7%) |  |

Values are median (Q1, Q3) for continuous variables and n (%) for categorical variables. N indicates the number of participants with available data. p-values are from Kruskal-Wallis rank-sum tests for continuous variables and from Pearson's χ² or Fisher's exact tests for categorical variables. Abbreviations: Aβ, amyloid-β; BVMT-R, Brief Visuospatial Memory Test-Revised; CDR, Clinical Dementia Rating; CKD-EPI, Chronic Kidney Disease Epidemiology Collaboration; CVLT-II, California Verbal Learning Test-Second Edition; eGFR, estimated glomerular filtration rate; MCI, mild cognitive impairment; PACC, Preclinical Alzheimer's Cognitive Composite; PASE, Physical Activity Scale for the Elderly; PET, positron emission tomography; PHQ-9, Patient Health Questionnaire-9; PiB, Pittsburgh compound B.

Baseline characteristics were generally comparable between the placebo group (n = 39) and the Li group (n = 41), except that the Li group exhibited higher estimated glomerular filtration rates and lower creatinine levels (Supplemental Table 1). Among the 75 participants with available Aβ-PET status, comparisons across treatment-by-PiB subgroups revealed nominal differences in MCI subtype, comorbidity burden, cognitive performance, and brain volumes. Within each treatment group, PiB-positive subgroups exhibited lower cognitive scores and reduced hippocampal volumes (Supplemental Table 2).

### Baseline differential protein levels by Li treatment assignment and Aβ-PET status

Figure 1 presents baseline differences in circulating protein levels by randomized treatment assignment and Aβ-PET status, adjusted for age and sex. Fold changes and p-values reported in the text are from the age- and sex-adjusted models shown in the volcano plots, whereas box-plot annotations show unadjusted, median-based fold changes and Wilcoxon rank-sum p-values. In comparisons by randomized treatment assignment, five proteins exhibited nominal differences between the Li and placebo groups after adjustment for age and sex (see Figure 1a): CSF2 (FC 0.79, p = 0.009), CALB2 (FC 0.84, p = 0.045), and S100B (FC 0.82, p = 0.049) were lower in the Li group, while IL10 (FC 1.48, p = 0.020) and MME (FC 2.12, p = 0.022) were higher. CX3CL1 was nominally lower in the Li group only in the unadjusted comparison (FC 0.88, p = 0.035) (Supplemental Table 3). None of the treatment-assignment differences remained significant after false discovery rate (FDR) correction, consistent with successful randomization. Consistent with these limited differences, single-protein ROC analyses showed only modest discrimination between treatment assignments, with AUC values ranging from 0.610 for CCL17 to 0.690 for CSF2 among the proteins displayed (Supplemental Figure 2a).

**Figure 1.**
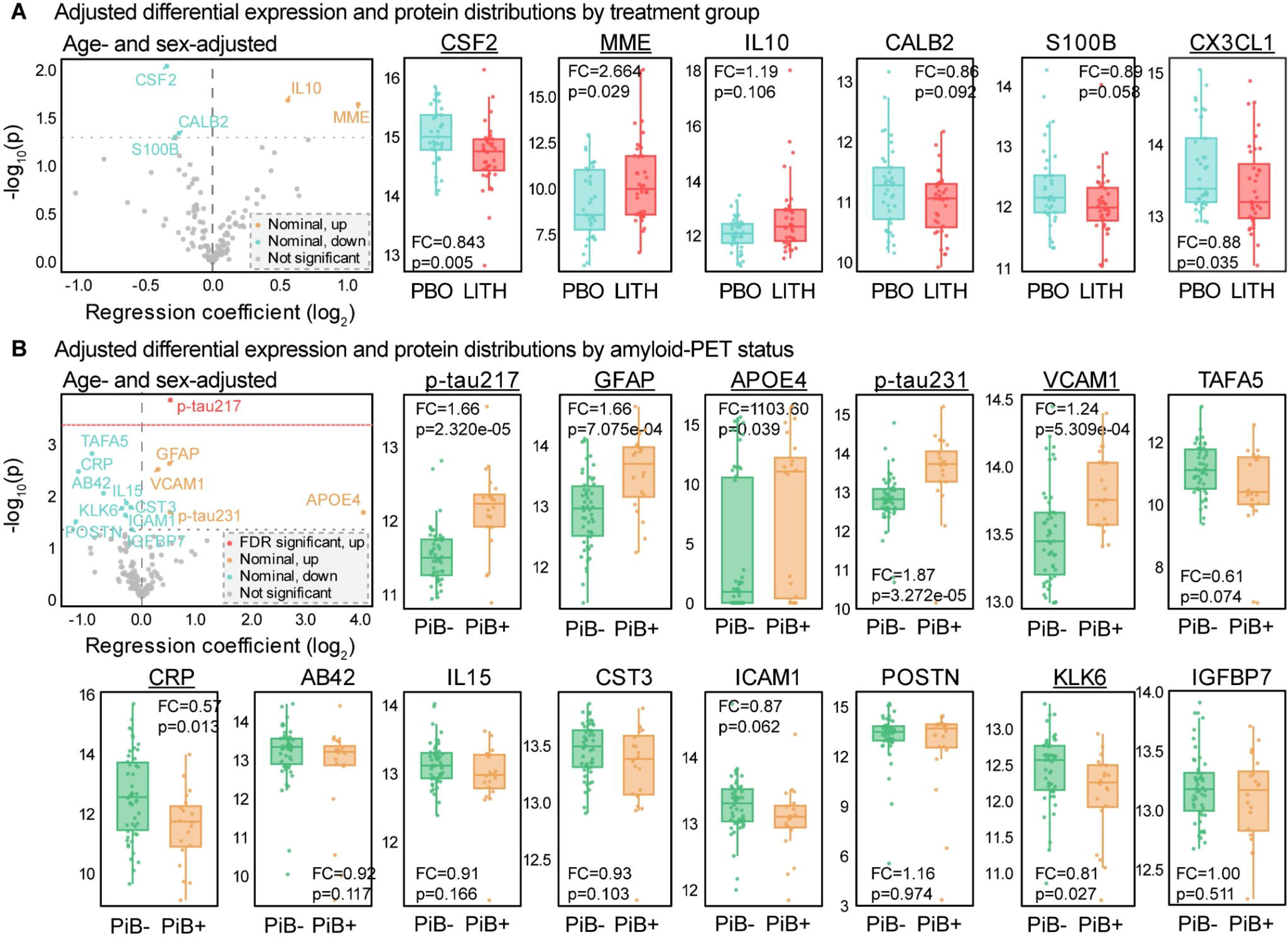
Baseline differential protein abundance by randomized treatment assignment and Aβ-PET status. **(A)** Volcano plot (left) and selected protein distributions (right) comparing participants assigned to Li (LITH) versus placebo (PBO). **(B)** Volcano plot (left) and selected protein distributions (right) compare Aβ-PET-positive (PiB+) and Aβ-PET-negative (PiB-) participants. Volcano plots display regression coefficients on the log_2_ scale (x-axis) and -log_10_ nominal p-values (y-axis) from age- and sex-adjusted limma linear regression models with empirical Bayes moderation. Positive coefficients correspond to higher protein levels in Li-assigned participants in panel **(A)** or PiB+ participants in panel **(B)**. Teal and orange indicate nominally lower and higher protein levels, respectively; red indicates an FDR-significant association, and grey indicates nonsignificant associations. Dotted grey horizontal lines represent p = 0.05, and the red horizontal line represents the Benjamini-Hochberg significance threshold at FDR = 0.05. Box plots present baseline protein levels in log_2_ NPQ units by treatment assignment (PBO, teal; LITH, red) or PiB status (PiB-, green; PiB+, orange). Boxes indicate medians and interquartile ranges; points represent individual participants. Annotations report median-based fold changes calculated on log_2_ scale and two-sided Wilcoxon rank-sum p-values. Underlined protein names indicate Wilcoxon p<0.05. Baseline measurements were obtained prior to Li treatment initiation; differences by treatment assignment do not reflect Li treatment effects. Abbreviations: FC, fold change; FDR, false discovery rate; Li/LITH, lithium; NPQ, NULISA Protein Quantification; PBO, placebo; PET, positron emission tomography; PiB, Pittsburgh compound B amyloid-PET status.

More proteins differed by Aβ-PET status (see Figure 1b). P-tau217 was elevated in PiB+ participants (FC 1.43, p = 1.58 × 10^-4^, FDR = 0.020) and was the only protein to remain significant after FDR correction. GFAP (FC 1.42, p = 0.003), VCAM1 (FC 1.23, p = 0.003), p-tau231 (FC 1.42, p = 0.023), and APOE4 (FC 16.40, p = 0.023) were nominally higher in PiB+ participants, while TAFA5 (FC 0.53, p = 0.002), CRP (FC 0.45, p = 0.004), AB42 (FC 0.62, p = 0.010), IL15 (FC 0.82, p = 0.015), CST3 (FC 0.87, p = 0.019), KLK6 (FC 0.78, p = 0.019), ICAM1 (FC 0.81, p = 0.026), POSTN (FC 0.43, p = 0.035), and IGFBP7 (FC 0.88, p = 0.0498) were present at nominally lower plasma levels in PiB+ participants (Supplemental Table 4). Higher APOE4 protein levels in PiB+ participants are consistent with the greater proportion of APOE ε4 carriers in this group (60.0% versus 28.0% among participants with baseline plasma; Fisher’s exact p = 0.016). TAFA5, AB42, IL15, CST3, ICAM1, POSTN, and IGFBP7 did not reach nominal significance in unadjusted comparisons (box plots in Figure 1b).

To assess baseline balance within the PiB randomization strata, we examined proteomic profiles of four subgroups stratified by PiB and treatment: placebo/PiB-, placebo/PiB+, Li/PiB-, and Li/PiB+ (see Supplemental Figure 1). Across these subgroups, we identified nominal differences in 25 proteins (p < 0.05; see Supplemental Figure 1a), of which p-tau217 and p-tau231 remained significant after FDR correction. After adjusting for age and sex and conducting pairwise comparisons, we observed that, within the PiB+ subgroup, participants receiving Li had higher levels of several proteins than those receiving placebo, including p-SNCA129, SNCA, ANXA5, RUVBL2, TARDBP, MDH1, CD40LG, PGK1, and oligomeric α-synuclein (all FDR < 0.05). In the PiB-subgroup, Li recipients had nominally lower levels of CALB2, S100B, NPTX2, PARK7, CSF2, CX3CL1, ANXA5, and CXCL8 (see Supplemental Figure 1b). Single-protein discrimination between treatment assignments was also greater within the PiB+ subgroup, with CD40LG showing the highest AUC (0.889), followed by p-SNCA129 (0.879), p-TDP43 409 (0.859), and TARDBP (0.859). In the PiB-subgroup, S100B had the highest AUC (0.747), followed by CSF2 (0.723) and VEGFD (0.700; Supplemental Figure 2a).

Comparisons by Aβ-PET status within each treatment assignment revealed additional differences. In the Li group, PiB+ participants exhibited nominally higher levels of RUVBL2, CXCL8, GOT1, PGK1, p-SNCA129, SNCA, IL18, and ANXA5, as well as lower TAFA5 levels. In the placebo group, PiB+ participants had higher levels of p-tau217 (FDR = 0.033) and nominally higher levels of APOE4, GFAP, VCAM1, and p-tau231, and lower levels of several proteins, including ANXA5, MDH1, CD40LG, TARDBP, p-SNCA129, SNCA, CRP, and SOD1 (Supplemental Figure 1b).

In summary, baseline protein differences were more pronounced by Aβ-PET status than by randomized treatment assignment, with only p-tau217 remaining significant after FDR correction in the overall PiB comparison. Differences between treatment assignments, including protein-level discrimination, reflect baseline variation before treatment initiation and should not be interpreted as effects of Li.

### Baseline differential protein abundance by amnestic MCI status

In age- and sex-adjusted comparisons of participants with aMCI and non-aMCI, eight proteins reached nominal significance (Figure 2; Supplemental Table 5). VCAM1 (FC 1.20, p = 0.009), BACE1 (FC 1.22, p = 0.010), KDR (FC 2.10, p = 0.021), and VGF (FC 2.04, p = 0.024) levels were elevated in aMCI participants. In contrast, CCL3 (FC 0.48, p = 6.01 × 10), TREM1 (FC 0.81, p = 0.015), AB42 (FC 0.68, p = 0.036), and CHI3L1 (FC 0.62, p = 0.036) were nominally reduced, with CCL3 showing the strongest association (FDR = 0.076). In unadjusted comparisons (box plots), VGF, TREM1, and CHI3L1 did not reach nominal significance. Conversely, p-tau217 was nominally higher in aMCI participants in the unadjusted comparison (FC 1.23, p = 0.026) but not after adjustment for age and sex (FC 1.16, p = 0.120).

**Figure 2.**
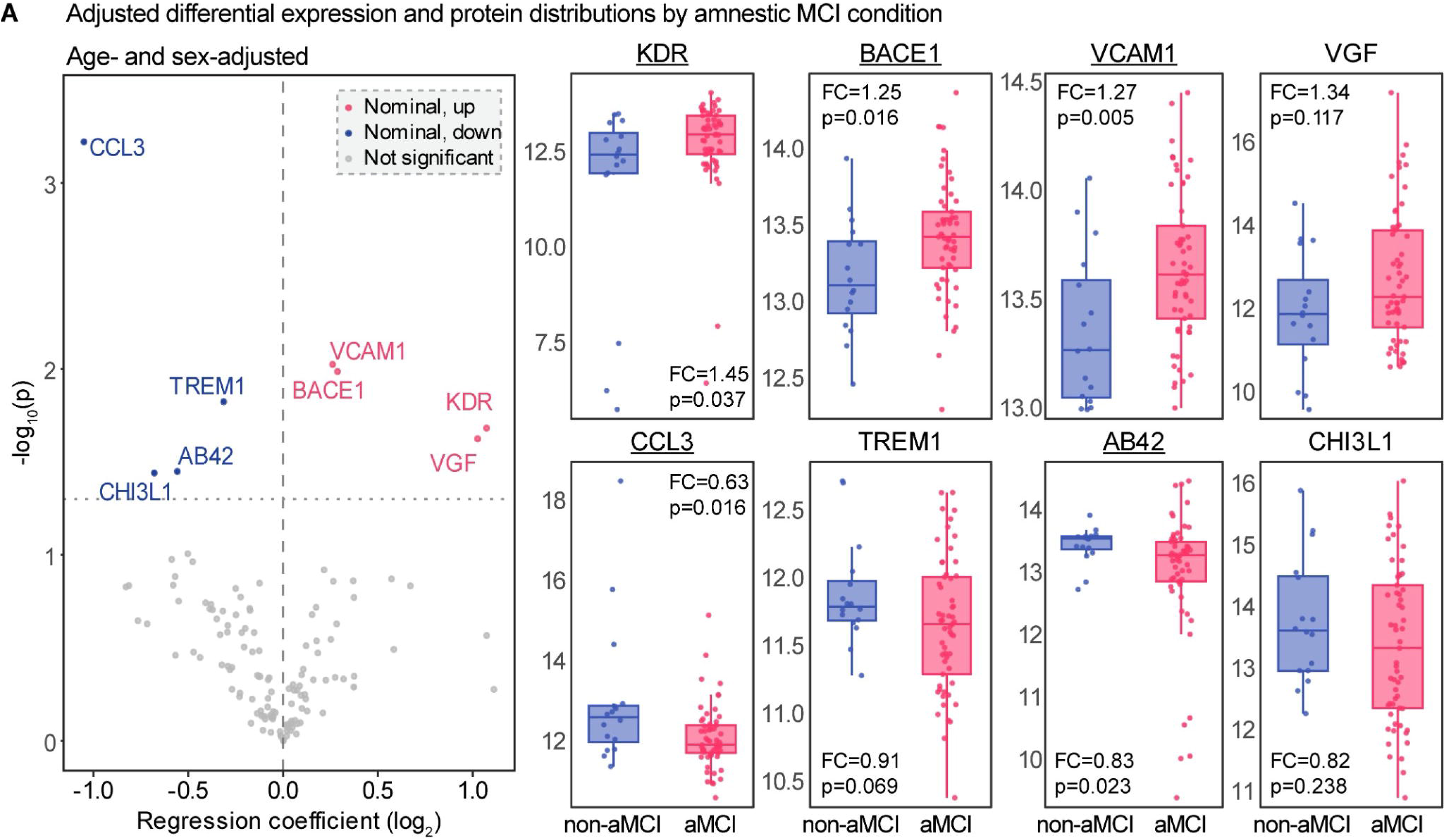
Baseline differential protein abundance by amnestic MCI status. The volcano plot (left) and selected protein distributions (right) compare participants with aMCI and non-aMCI. The volcano plot presents regression coefficients on the log_2_ scale (x-axis) and - log_10_ nominal p-values (y-axis) derived from age- and sex-adjusted limma linear regression models with empirical Bayes moderation. Positive coefficients correspond to higher protein levels in aMCI participants. Pink and blue indicate nominally higher and lower protein levels, respectively (p<0.05), grey represents nonsignificant differences. The dotted horizontal line indicates p = 0.05. Box plots display baseline protein levels in log_2_ NPQ units for eight selected proteins for non-aMCI (blue) and aMCI (pink) participants. Boxes indicate medians and interquartile ranges; points represent individual participants. Annotations report median-based fold changes on the log_2_ scale and unadjusted two-sided Wilcoxon rank-sum p values. Underlined protein names indicate Wilcoxon p < 0.05. No differential-abundance association remained significant after FDR correction. Abbreviations: aMCI, amnestic mild cognitive impairment; FC, fold change; FDR, false discovery rate; MCI, mild cognitive impairment; NPQ, NULISA Protein Quantification.

ROC analyses for individual proteins indicated modest discrimination between aMCI and non-aMCI participants, with VCAM1 having the highest AUC (0.732), followed by BACE1 and CCL3 (both 0.698; Supplemental Figure 2b). Among participants with aMCI, p-tau217 provided the strongest discrimination of Aβ-PET status (AUC = 0.808), followed by p-tau231 (AUC = 0.803) and GFAP (AUC = 0.744; Supplemental Figure 2c).

Overall, baseline protein differences between aMCI and non-aMCI participants were modest and did not survive multiple-testing correction. Within participants with aMCI, phosphorylated tau markers showed the strongest discrimination of Aβ-PET status among the proteins examined.

### Li treatment was associated with nominal differences in plasma protein trajectories over a two-year period

Longitudinal analyses evaluated whether plasma protein trajectories differed between participants randomized to receive daily oral Li carbonate or placebo from baseline (T1) through Year 1 (T2) and Year 2 (T3) in the LATTICE trial (Figure 3). Treatment-by-time interaction terms were used to assess whether changes across visits differed between the two groups. In the overall cohort, CALB2 and TREM2 exhibited nominally significant treatment-by-time interactions (β□=□0.125 [SE, 0.057], p□=□0.031 and β□=□0.093 [SE, 0.043], p□=□0.034, respectively; β denotes the Li-placebo difference in change in log_2_ NPQ per visit interval; Figure 3a). For these proteins, participants treated with Li demonstrated a relative increase or a smaller decline over time compared with those assigned to placebo, corresponding to model-implied relative differences in change from T1 to T3 of 1.19-fold for CALB2 and 1.14-fold for TREM2. However, neither interaction remained significant after FDR correction (both FDR = 0.988). Within the aMCI subgroup (Figure 3b), Li treatment-by-time interactions were larger for both CALB2 (β□=□0.170 [SE, 0.067], Tx×time p□=□0.013, FDR□=□0.836) and TREM2 (β□=□0.129 [SE, 0.050], Tx×time p□=□0.013, FDR□=□0.836) and were also observed for AB40 (β□=□0.233 [SE, 0.097], Tx×time p□=□0.020, FDR□=□0.867) and TEK (β□= □.112 [SE, 0.055], Tx×time p = 0.046, FDR = 0.882). Similar to the full cohort, these proteins also showed a relative increase or a smaller decline across visits in the Li group compared with placebo (model-implied relative differences in change from T1 to T3: 1.27-fold for CALB2, 1.20-fold for TREM2, 1.38-fold for AB40, and 1.17-fold for TEK). Overall, Li assignment was associated with nominal differences in the two-year trajectories of CALB2, TREM2, AB40, and TEK plasma proteins. However, these findings did not remain significant after correction for multiple testing, providing limited evidence that Li altered protein trajectories.

**Figure 3.**
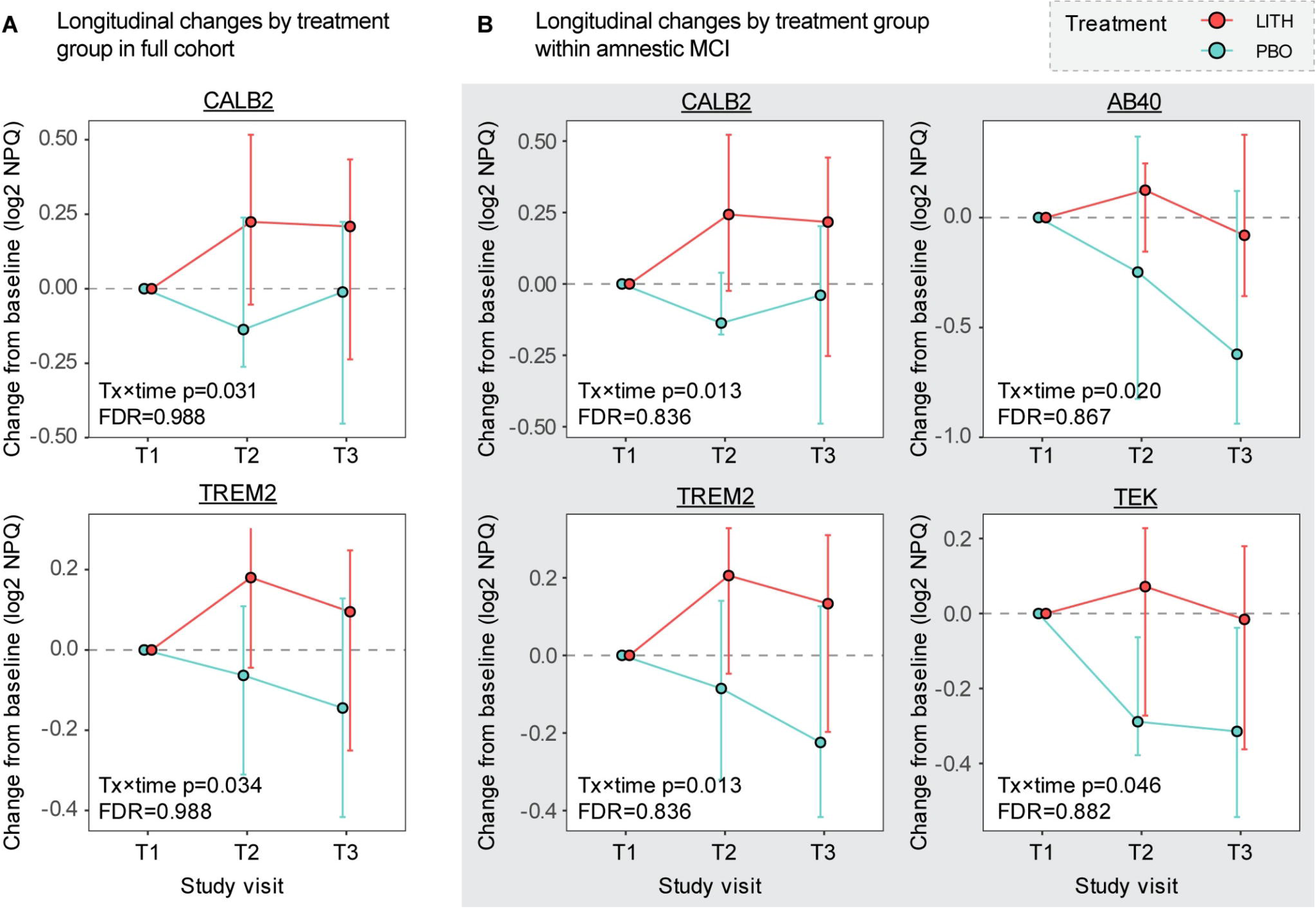
Longitudinal plasma protein changes by randomized treatment assignment. **(A)** Full cohort: CALB2 and TREM2. **(B)** Baseline amnestic MCI (aMCI) subgroup: CALB2, TREM2, AB40, TEK. Proteins shown are those with a nominally significant treatment × time interaction (p<0.05, underlined protein names) in the corresponding population. For each participant, the baseline (T1) value was subtracted from the values at Year 1 (T2) and Year 2 (T3), aligning all trajectories start at zero (dashed line, no change). Values are differences in log_2_ NPQ and therefore represent log_2_ fold change from baseline. Circles indicate group medians, and error bars show interquartile ranges, for placebo (PBO, teal) and Li (LITH, red). Treatment × time interactions test whether linear changes over time differ between treatment groups, using linear mixed-effects models fitted to all available samples, adjusted for baseline age and sex, and including participant-level random effects. Each panel reports the nominal interaction p-value and its Benjamini-Hochberg FDR-adjusted value across all 127 proteins within the corresponding population. No interaction remained significant after FDR correction. Abbreviations: aMCI, amnestic mild cognitive impairment; FDR, false discovery rate; LITH, lithium; MCI, mild cognitive impairment; NPQ, NULISA Protein Quantification; PBO, placebo; Tx, treatment.

### GDF15 and p-tau217 demonstrated distinct associations with neurodegeneration and clinical progression

We next examined whether baseline plasma protein levels were associated with subsequent cognitive, functional, and imaging trajectories and whether they modified the effect of Li on these trajectories, in both the full cohort and the aMCI subgroup. All 127 proteins were evaluated using three approaches: standalone models evaluated whether each protein was associated with longitudinal change; add-on models determined whether these associations persisted after accounting for treatment-by-time effects. Whereas these two prognostic models examined associations between baseline protein levels and outcome trajectories, predictive models assessed whether the effect of Li over time differed by baseline protein levels. Nine proteins (p-tau217, GDF15, IL-16, ANXA5, UCHL1, POSTN, CHIT1, TIMP3, and PGK1) each showed at least one association that remained significant after FDR correction and are presented in Figure 4 (p-tau217 and GDF15) and Supplemental Figure 3 (the remaining seven proteins).

**Figure 4.**
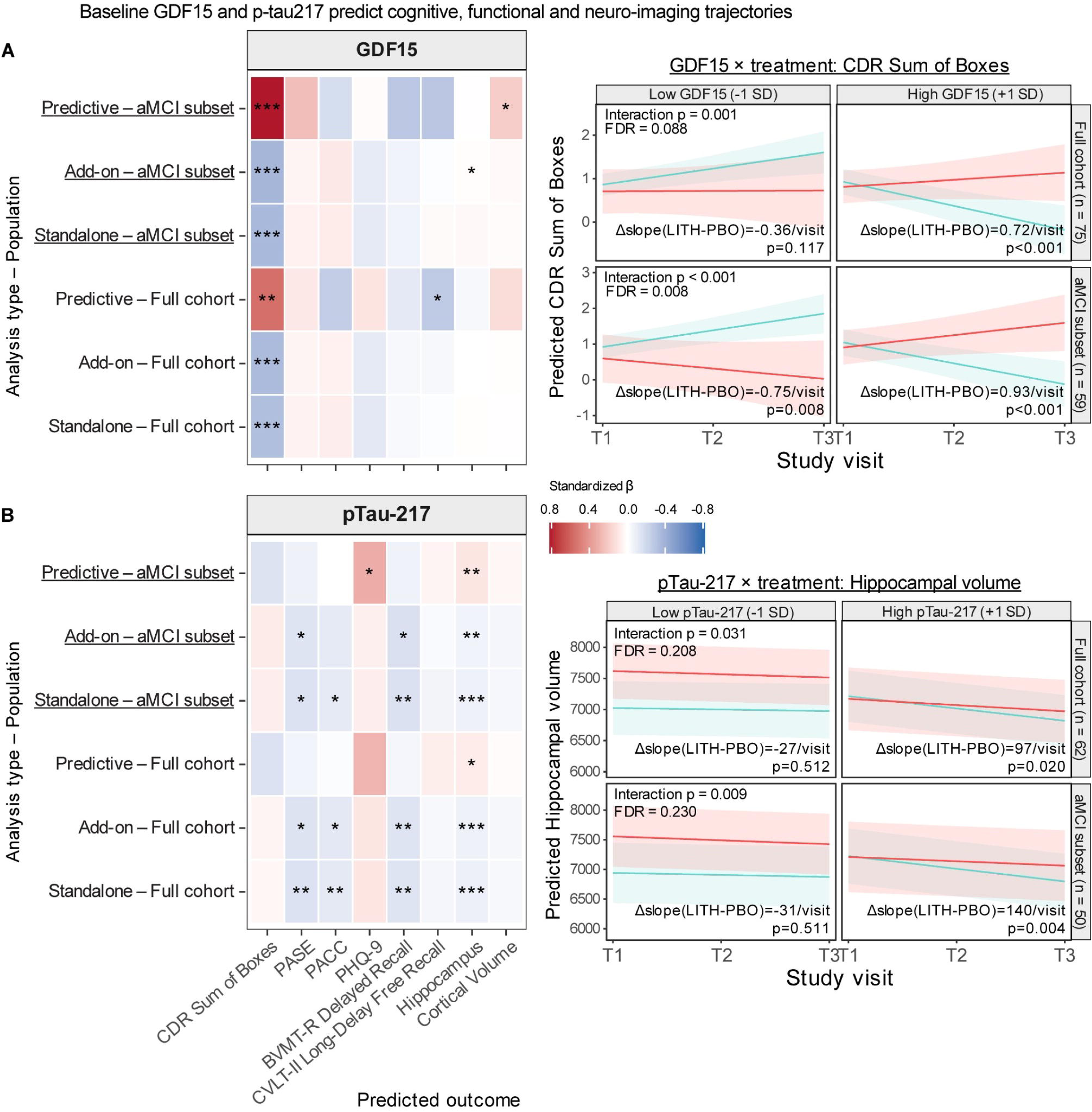
Associations of baseline GDF15 and p-tau217 with cognitive, functional and neuro-imaging trajectories. **(A)** GDF15. **(B)** p-tau217. Left: Heatmaps show standardized β coefficients relating baseline marker levels to changes in eight outcomes. Outcomes are ordered as functional (CDR Sum of Boxes, PASE), cognitive (PACC, PHQ-9, BVMT-R Delayed Recall, CVLT-II Long-Delay Free Recall) and MRI imaging measures (hippocampal volume, cortical volume). Each marker was evaluated using three linear mixed-effects models adjusted for baseline age, sex, and PiB status and including participant-level random effects. Standalone models tested the baseline marker × time interaction without treatment terms. Add-on models used a likelihood-ratio test to assess whether jointly adding the baseline marker main effect and marker × time interaction improved a model containing treatment × time. Predictive models used a likelihood-ratio test of the baseline marker × treatment × time interaction to assess whether the marker-trajectory association differed between treatment groups. Models were fitted separately in the full cohort and the baseline aMCI subset (underlined rows). Baseline marker levels were z-scored within each analytic sample. Fill indicates standardized β, expressed as SD change in outcome per year per 1-SD higher baseline marker level; for Predictive rows, β represents the difference in this association between lithium and placebo. Both heatmaps share a common color scale, with red indicating positive coefficients and blue indicating negative coefficients. Clinical interpretation depends on the outcome and model type. Stars indicate nominal p values: * p < 0.05, ** p < 0.01, and *** p < 0.001. Right: model-predicted trajectories of CDR Sum of Boxes for GDF15 (a) and hippocampal volume for p-tau217 (b), evaluated at low (-1 SD) and high (+1 SD) baseline marker levels. Lines indicate predicted means for placebo (teal) and lithium (red); shaded bands show 95% confidence intervals. Predictions are shown separately for the full cohort and aMCI subset, with sample sizes reported in the row labels. Predictions hold age at its mean and average over sex and PiB status according to their proportions in the analytic sample. Annotations report the three-way interaction p value and FDR-adjusted value, together with the lithium-minus-placebo slope difference (Δslope, per visit interval) and its nominal p value at each marker level. Abbreviations: aMCI, amnestic mild cognitive impairment; BVMT-R, Brief Visuospatial Memory Test-Revised; CDR, Clinical Dementia Rating; CVLT-II, California Verbal Learning Test-Second Edition; FDR, false discovery rate; GDF15, growth differentiation factor 15; LITH, lithium; PACC, Preclinical Alzheimer’s Cognitive Composite; PASE, Physical Activity Scale for the Elderly; PBO, placebo; PHQ-9, Patient Health Questionnaire-9; PiB, Pittsburgh compound B; p-tau217, tau phosphorylated at threonine 217; SD, standard deviation.

GDF15 demonstrated its strongest associations with CDR Sum of Boxes trajectories in both standalone and add-on models for the full cohort and the aMCI subgroup (all p < 0.001; all FDR ≤ 0.014), with higher baseline GDF15 associated with a more negative CDR Sum of Boxes slope (standardized β, -0.33 to -0.37). It also exhibited treatment-effect modification for this outcome in the full cohort (standardized β = 0.55, p = 0.001, FDR = 0.088) and in the aMCI subgroup (standardized β = 0.82, p < 0.001, FDR = 0.008), indicating that the association between baseline GDF15 and CDR Sum of Boxes change was more positive in the Li group than in the placebo group. In model-predicted trajectories (Figure 4a, right), the Li-placebo difference in CDR Sum of Boxes slope was positive at high baseline GDF15 (+1 SD; Δslope = 0.72 per visit, p < 0.001 in the full cohort; 0.93 per visit, p < 0.001 in the aMCI subgroup) and negative or null at low baseline GDF15 (-1 SD; -0.36 per visit, p = 0.117; and - 0.75 per visit, p = 0.008, respectively). In the aMCI subgroup, this treatment-effect modification remained significant after FDR correction. Additional nominal findings included an association with hippocampal volume in the aMCI add-on model, as well as treatment-effect modification for cortical volume in the aMCI subgroup and for CVLT-II Long-Delay Free Recall in the full cohort (all p < 0.05).

As expected, p-tau217 showed its strongest associations with hippocampal volume trajectories. In both standalone and add-on models, these associations were observed in the full cohort (p < 0.001; FDR = 0.002 and 0.019, respectively) and in the aMCI subgroup (p < 0.001 and p < 0.01; FDR = 0.022 and 0.19, respectively), with higher baseline p-tau217 associated with greater hippocampal volume loss. Treatment-effect modification for hippocampal volume was observed in the full cohort (standardized β = 0.08, p < 0.05) and in the aMCI subgroup (standardized β = 0.10, p < 0.01), indicating a less negative association between baseline p-tau217 and hippocampal volume change in the Li group than in the placebo group; neither remained significant after FDR correction (FDR = 0.21 and 0.23, respectively). In model-predicted trajectories (Figure 4b, right), the Li-placebo difference in hippocampal volume slope was positive at high baseline p-tau217 (+1 SD; Δslope = 97 mm^3^ per visit, p = 0.020 in the full cohort; 140 mm^3^ per visit, p = 0.004 in the aMCI subgroup) but not at low baseline p-tau217 (-1 SD; -27 mm^3^ per visit, p = 0.512; and -31 mm^3^ per visit, p = 0.511, respectively). Beyond hippocampal volume, p-tau217 was associated with PASE and BVMT-R Delayed Recall trajectories in both models and across both populations. Associations with PACC reached nominal significance in both full-cohort models but only in the standalone aMCI model. Additionally, p-tau217 demonstrated nominal treatment-effect modification for PHQ-9 trajectories in both populations (standardized β = 0.29 and 0.31; p < 0.05; both FDR > 0.97).

Seven additional proteins demonstrated outcome-specific associations (see Supplemental Figure 3). In both the full cohort and the aMCI subgroup, ANXA5 and UCHL1 exhibited nominal treatment-effect modification for PACC trajectories, while IL-16 showed treatment-effect modification for CDR Sum of Boxes; in the aMCI subgroup, these effect modifications remained significant after FDR correction (FDR = 0.034, 0.014, and 0.011 for ANXA5, UCHL1, and IL-16, respectively). POSTN was associated with CDR Sum of Boxes trajectories in both standalone and add-on models (all FDR ≤ 0.024) and nominally demonstrated treatment-effect modification for BVMT-R Delayed Recall. CHIT1 was associated with PACC trajectories and TIMP3 with BVMT-R Delayed Recall trajectories in the respective models (FDR < 0.05 in the standalone and add-on models in the aMCI subgroup), whereas PGK1 was associated with cortical volume in the add-on models for both groups (FDR = 0.025 and 0.023, respectively), as was ANXA5 (FDR = 0.001 and < 0.001, respectively).

In summary, GDF15 showed consistent associations with CDR Sum of Boxes trajectories and may affect Li treatment response in both the full cohort and the aMCI subgroup. p-tau217 was most consistently linked to hippocampal volume trajectories, while other proteins displayed outcome-specific associations. Associations that did not survive FDR correction should be considered exploratory.

## Discussion

Clinical studies have assessed Li’s neuroprotective potential in MCI and AD, as well as its impact on clinical and biological outcomes ^24,29^. In this secondary proteomic analysis of the LATTICE trial, baseline proteomic variation was more strongly associated with Aβ status than with treatment assignment, with p-tau217 showing the strongest association with Aβ-PET positivity. Longitudinal analyses identified nominal Li-associated changes in CALB2 and TREM2, with an additional association for Aβ40 in the aMCI subgroup, although none remained significant after multiple-testing correction. Growth differentiation factor (GDF)15 showed the most consistent associations with clinical progression and potential treatment-response heterogeneity, whereas p-tau217 was more closely associated with hippocampal-volume trajectories.

The baseline proteomic analysis demonstrated Aβ pathology was a major cause of molecular heterogeneity in the cohort. As expected, p-tau217 showed the strongest association with Aβ-PET positivity and was the only protein to remain significant after FDR correction. GFAP, VCAM1, p-tau231, and APOE4 levels were nominally higher in PiB-positive participants, while CRP and KLK6 were nominally lower. About the MCI subtypes, aMCI had nominally higher levels of VCAM1, BACE1, and KDR and nominally lower levels of CCL3 and Aβ42, although none of these remained significant after correction for multiple testing. These proteins are involved in several biological processes relevant to AD, including as Aβ and tau biology (Aβ42, BACE1, p-tau217, p-tau231, and APOE4), astroglial and inflammatory signalling (GFAP, CCL3, and CRP), vascular-endothelial signalling (VCAM1 and KDR), and extracellular proteolysis and tissue remodelling (KLK6). Apart from the anticipated p-tau217 signal, the overall baseline proteomic profile shows that Aβ status and MCI phenotype are associated with differences in several biological processes. Since these differences were observed before treatment, they represent baseline variations related to the disease.

Longitudinal analyses identified nominal treatment-by-time interactions for CALB2 and TREM2 in the full cohort, and for CALB2, TREM2 and Aβ40 in the aMCI subgroup. For each protein, the Li group showed a relative increase or smaller decline over time compared with placebo. CALB2 encodes calretinin, a calcium-binding protein whose buffering function influences neuronal excitability in experimental models ^31^. TREM2 regulates microglial phagocytosis and inflammatory responses, including clearance of apoptotic neurons ^32^. Aβ40 is relevant to amyloid metabolism ^33^. Together with Li’s well-known inhibition of GSK-3α and GSK-3β, enzymes involved in amyloid processing and tau phosphorylation, these observations provide a rationale for studying links among calcium homeostasis, microglial responses, and Li exposure. However, plasma protein trajectories do not directly demonstrate changes in neuronal calcium buffering, microglial activity, or central target engagement. However, none of these interactions remained significant after FDR correction and should therefore be interpreted as exploratory molecular signals rather than definitive pharmacodynamic effects of Li.

Although LATTICE showed only limited cognitive and neuroimaging treatment signals ^29^, GDF15 emerged as the most consistent proteomic marker of clinical progression and potential heterogeneity in treatment response. Baseline GDF15 was strongly associated with longitudinal CDR Sum of Boxes (CDR-SB) trajectories in both the full cohort and the aMCI subgroup, across standalone and add-on models, and modified treatment effects for CDR-SB. We also observed additional associations with hippocampal volume in the aMCI add-on model, cortical volume in the aMCI predictive model, and CVLT-II Long-Delay Free Recall in the full-cohort predictive model. These results align with accumulating evidence that links elevated GDF15 concentrations to cognitive decline and increased dementia risk ^34^. Large prospective cohorts, such as the UK Biobank, ARIC, and the Framingham Offspring Study, have demonstrated associations between circulating GDF15 and subsequent dementia ^35–37^. Elevated peripheral GDF15 has also been linked to cerebral small-vessel disease and white-matter hyperintensities ^38^. Additionally, cerebrospinal fluid GDF15 has been associated with p-tau181, sTREM2, GFAP, and neurofilament light ^39^. On the other hand, p-tau217 in this group was most consistently related to the trajectories of hippocampal volume. Collectively, these findings indicate that GDF15 and p-tau217 capture complementary aspects of disease progression. GDF15 reflects broader systemic, vascular, inflammatory, metabolic, and cellular stress processes, whereas p-tau217 is more closely associated with Alzheimer’s disease-related Aβ pathology and hippocampal structural changes. Given that GDF15 is not specific to Alzheimer’s disease, its potential utility as a prognostic or treatment-response biomarker requires validation in larger, adequately powered cohorts.

In addition to GDF15 and p-tau217, seven other proteins demonstrated exploratory associations with cognitive, functional, and imaging outcomes. Regarding functional outcomes, IL-16 exhibited nominal treatment-effect modification for CDR-SB, while POSTN was associated with CDR-SB trajectories. For cognitive outcomes, ANXA5 and UCHL1 showed treatment-effect modification for PACC trajectories; CHIT1 was associated with PACC; and POSTN and TIMP3 were associated with BVMT-R delayed recall. For imaging outcomes, PGK1 was associated with cortical-volume trajectories. These proteins represent diverse pathways, including immune and inflammatory signaling (IL-16, CHIT1), extracellular matrix and tissue remodeling (POSTN, TIMP3), cellular stress and membrane biology (ANXA5), neuronal protein homeostasis (UCHL1), and energy metabolism (PGK1). These outcome-specific associations provide additional candidate molecular signals but should be interpreted cautiously given their nominal significance.

This study has several limitations, including modest sample size, limited subgroup power, multiple nominal associations, no independent replication, and indirect measurement of plasma biomarkers. The longitudinal and treatment-modification results are hypothesis generating. The two-year follow-up period limits evaluation of long-term proteomic trajectories and clinical outcomes. Additionally, the absence of plasma lithium concentration measurements prevents analysis of associations between lithium exposure, proteomic changes, and clinical outcomes.

In conclusion, plasma proteomics identified GDF15 as a candidate marker for clinical progression and Li treatment response in MCI, while p-tau217 demonstrated a stronger association with Aβ pathology and hippocampal alterations. Exploratory findings indicate that Li may affect CALB2, TREM2, and Aβ40, suggesting molecular effects across neuronal, microglial, and Aβ-related pathways.

## Data availability

All relevant data that support this study are provided as supplementary files or available to any interested researchers upon reasonable request to the corresponding author

## Acknowledgements

This section is not mandatory.

## Abbreviations

Aβ: amyloid-β
AD: Alzheimer’s disease
aMCI: amnestic mild cognitive impairment
BVMT-R: Brief Visuospatial Memory Test-Revised
CVLT-II: California Verbal
GDF15: growth differentiation factor 15
LATTICE: Lithium as a Treatment to Prevent Impairment of Cognition in Elders
NULISA: nucleic acid-linked immuno-sandwich assay
PACC: Preclinical Alzheimer Cognitive Composite

---

**Supplemental Figure 1. Baseline protein abundance by randomized treatment assignment and Aβ-PET subgroup.**

**(A)** Box plots present baseline protein levels in log_2_-normalized NPQ units for 25 proteins exhibiting nominal differences across four subgroups: placebo/PiB-, placebo/PiB+, Li/PiB-, and Li/PiB+ in unadjusted Kruskal-Wallis tests (p<0.05): PBO/PiB-(teal), PBO/PiB+ (green), LITH/PiB-(orange), and LITH/PiB+ (red). Protein levels are expressed in log_2_ NPQ units. Boxes indicate medians and interquartile ranges; points represent individual participants. Brackets identify nominally significant pairwise comparisons from unadjusted two-sided Wilcoxon rank-sum tests. Annotations report median-based fold changes and corresponding unadjusted p-values with nominal significance. **(B)** Age- and sex-adjusted volcano plots compare Li versus placebo within PiB+ participants (upper left) and PiB-participants (upper right), and PiB+ versus PiB-participants within the Li-assigned group (lower left) and placebo-assigned group (lower right). Estimates are from limma linear models with empirical Bayes moderation. The x-axis represents regression coefficients on the log_2_ scale, and the y-axis represents -log_10_ nominal p-values. Orange and teal indicate nominally higher and lower levels, respectively (p < 0.05); red indicates FDR-significant higher levels, and gray indicates nonsignificant differences. Dotted horizontal lines indicate p = 0.05, and dashed vertical lines indicate a coefficient of zero. Baseline samples were collected before lithium treatment initiation; differences by randomized assignment therefore do not represent treatment effects. Abbreviations: Aβ, amyloid-β; FC, fold change; FDR, false discovery rate; LITH, lithium; NPQ, NULISA Protein Quantification; PBO, placebo; PET, positron emission tomography; PiB, Pittsburgh compound B.

**Supplemental Figure 2. Discrimination of baseline participant groups using individual plasma proteins.** Receiver operating characteristic (ROC) curves are shown for the ten proteins with highest observed area under the curve (AUC) in each comparison. AUC values are listed in the panel legends, with underlining identifies the protein with the highest AUC. **(A)** Randomized treatment assignment (Li versus placebo) in the full cohort (left), PiB+ subgroup (upper right), and PiB-subgroup (lower right). **(B)** Amnestic versus non-amnestic MCI. **(C)** PiB+ versus PiB-status among participants with amnestic MCI. Dashed diagonal lines indicate chance discrimination (AUC = 0.5). Discrimination between randomized treatment groups reflects baseline differences and does not represent a lithium treatment effect. Abbreviations: AUC, area under the curve; MCI, mild cognitive impairment; PiB, Pittsburgh compound B; ROC, receiver operating characteristic.

**Supplemental Figure 3 Associations of seven additional baseline plasma proteins with cognitive, functional and neuro-imaging trajectories.**

Heatmaps display standardized β coefficients relating baseline plasma IL-16, ANXA5, UCHL1, POSTN, CHIT1, TIMP3, and PGK1 in eight outcomes. Outcomes are ordered as functional (CDR Sum of Boxes, PASE), cognitive (PACC, PHQ-9, BVMT-R Delayed Recall, CVLT-II Long-Delay Free Recall) and MRI imaging measures (hippocampal volume, cortical volume). Each marker was evaluated using three linear mixed-effects models adjusted for baseline age, sex, and PiB status and including participant-level random effects. Standalone models tested the baseline marker × time interaction without treatment terms. Add-on models used a likelihood-ratio test to assess whether jointly adding the baseline marker main effect and marker × time interaction improved a model containing treatment × time. Predictive models used a likelihood-ratio test of the baseline marker × treatment × time interaction to assess whether the marker– trajectory association differed between treatment groups. Models were fitted separately in the full cohort and the baseline aMCI subset (underlined rows). Baseline marker levels were z-scored within each analytic sample. Fill indicates standardized β, expressed as SD change in outcome per year per 1-SD higher baseline marker level; for Predictive rows, β represents the difference in this association between lithium and placebo. All panels share a common color scale, with red indicating positive coefficients and blue indicating negative coefficients. Clinical interpretation depends on the outcome and model type; these colors do not uniformly indicate worsening or protection. Stars indicate nominal p values: * p < 0.05, ** p < 0.01, and *** p < 0.001. Abbreviations: aMCI, amnestic mild cognitive impairment; ANXA5, annexin A5; BVMT-R, Brief Visuospatial Memory Test–Revised; CDR, Clinical Dementia Rating; CHIT1, chitotriosidase 1; CVLT-II, California Verbal Learning Test–Second Edition; IL-16, interleukin-16; PACC, Preclinical Alzheimer’s Cognitive Composite; PASE, Physical Activity Scale for the Elderly; PGK1, phosphoglycerate kinase 1; PHQ-9, Patient Health Questionnaire-9; PiB, Pittsburgh compound B; POSTN, periostin; TIMP3, tissue inhibitor of metalloproteinases 3; UCHL1, ubiquitin C-terminal hydrolase L1.

**Supplemental Table 1.** Demographic and clinical characteristics by randomized treatment assignment.

| **Characteristic** | **N** | **Overall**<br>**N = 80** | **Placebo**<br>**N = 39** | **Lithium**<br>**N = 41** | **p-value** |
|---|---|---|---|---|---|
| **Age**, years | 80 | 71.5 (66.5, 78.2) | 70.2 (65.7, 78.4) | 71.5 (66.9, 78.0) | 0.567 |
| **Education**, years | 80 | 16.0 (14.0, 18.0) | 16.0 (16.0, 18.0) | 16.0 (14.0, 18.0) | 0.086 |
| **Sex** | 80 |  |  |  | 0.978 |
| Female |  | 45 (56.3%) | 22 (56.4%) | 23 (56.1%) |  |
| Male |  | 35 (43.8%) | 17 (43.6%) | 18 (43.9%) |  |
| **Race** | 80 |  |  |  | 0.365 |
| Asian |  | 1 (1.3%) | 1 (2.6%) | 0 (0.0%) |  |
| Black or African American |  | 8 (10.0%) | 5 (12.8%) | 3 (7.3%) |  |
| White |  | 71 (88.8%) | 33 (84.6%) | 38 (92.7%) |  |
| **Ethnicity** | 80 |  |  |  | 0.487 |
| Not Hispanic or Latino |  | 79 (98.8%) | 38 (97.4%) | 41 (100.0%) |  |
| Hispanic or Latino |  | 1 (1.3%) | 1 (2.6%) | 0 (0.0%) |  |
| **Aβ-PET status** | 80 |  |  |  | 1.000 |
| PiB- |  | 54 (67.5%) | 27 (69.2%) | 27 (65.9%) |  |
| PiB+ |  | 21 (26.3%) | 10 (25.6%) | 11 (26.8%) |  |
| Unknown |  | 5 (6.3%) | 2 (5.1%) | 3 (7.3%) |  |
| **MCI subtype** | 80 |  |  |  | 0.233 |
| Amnestic MCI |  | 62 (77.5%) | 28 (71.8%) | 34 (82.9%) |  |
| Non-amnestic MCI |  | 18 (22.5%) | 11 (28.2%) | 7 (17.1%) |  |
| **Burden score** | 80 | 1.5 (0.0, 4.0) | 1.0 (0.0, 4.0) | 2.0 (1.0, 3.1) | 0.349 |
| **eGFR (CKD-EPI 2021)** | 80 | 84.2 (71.4, 91.9) | 77.0 (68.8, 90.2) | 85.8 (76.8, 92.5) | 0.026 |
| **Serum creatinine**, mg/dL | 80 | 0.9 (0.8, 1.0) | 0.9 (0.8, 1.0) | 0.8 (0.7, 1.0) | 0.022 |
| **Number of medications** | 80 | 6.0 (4.0, 9.0) | 5.0 (4.0, 9.0) | 6.0 (5.0, 10.0) | 0.244 |
| **CDR Global score** | 80 |  |  |  | 0.949 |
| 0 |  | 29 (36.3%) | 14 (35.9%) | 15 (36.6%) |  |
| 0.5 |  | 51 (63.8%) | 25 (64.1%) | 26 (63.4%) |  |
| **CDR Sum of Boxes** | 80 | 0.5 (0.3, 1.0) | 0.5 (0.0, 1.5) | 0.5 (0.5, 1.0) | 0.917 |
| **PHQ-9 total score** | 80 | 3.0 (1.0, 5.0) | 3.0 (1.0, 5.0) | 3.0 (2.0, 7.0) | 0.506 |
| **PASE total score** | 79 | 92.1 (61.8, 133.3) | 69.6 (59.3, 133.3) | 111.2 (69.6, 130.3) | 0.102 |
| **PACC** | 77 | 0.8 (-2.3, 2.0) | 0.8 (-2.2, 2.3) | 0.7 (-2.7, 1.9) | 0.420 |
| **Hippocampal volume**, mm<sup>3</sup> | 62 | 7,392.4 (6,630.7, 7,749.3) | 7,337.0 (6,472.8, 7,669.9) | 7,567.2 (6,914.3, 7,970.3) | 0.140 |
| **BVMT-R Delayed Recall** | 79 | 7.0 (4.0, 9.0) | 7.0 (4.0, 9.0) | 6.5 (4.0, 8.5) | 0.594 |
| **CVLT-II Long-Delay Free Recall** | 79 | 8.0 (6.0, 10.0) | 7.0 (5.0, 11.0) | 8.0 (6.0, 10.0) | 0.809 |
| **Cortical volume**, mm<sup>3</sup> | 62 | 408,790.2 (397,107.8, 428,692.6) | 404,250.2 (397,269.0, 417,455.5) | 420,939.1 (397,107.8, 437,297.5) | 0.066 |

Values are median (Q1, Q3) for continuous variables and n (%) for categorical variables. N indicates the number of participants with available data. p-values are from Kruskal-Wallis rank-sum tests for continuous variables and from Pearson’s χ<sup>2</sup> or Fisher’s exact tests for categorical variables. Abbreviations: Aβ, amyloid-β; BVMT-R, Brief Visuospatial Memory Test-Revised; CDR, Clinical Dementia Rating; CKD-EPI, Chronic Kidney Disease Epidemiology Collaboration; CVLT-II, California Verbal Learning Test-Second Edition; eGFR, estimated glomerular filtration rate; MCI, mild cognitive impairment; PACC, Preclinical Alzheimer’s Cognitive Composite; PASE, Physical Activity Scale for the Elderly; PET, positron emission tomography; PHQ-9, Patient Health Questionnaire-9; PiB, Pittsburgh compound B.

**Supplemental Table 2.** Demographic and clinical characteristics by randomized treatment assignment and Aβ-PET status.

| Characteristic | N | Overall<br>N = 75 | Placebo, PiB−<br>N = 27 | Placebo, PiB+<br>N = 10 | Lithium, PiB−<br>N = 27 | Lithium, PiB+<br>N = 11 | p-value |
|---|---|---|---|---|---|---|---|
| **Age**, years | 75 | 71.5 (66.4, 78.4) | 69.0 (63.1, 77.3) | 73.7 (66.8, 79.2) | 70.1 (64.8, 74.5) | 78.5 (71.5, 86.8) | 0.067 |
| **Education**, years | 75 | 16.0 (14.0, 18.0) | 16.0 (15.0, 18.0) | 17.0 (16.0, 18.0) | 16.0 (13.0, 18.0) | 17.0 (14.0, 18.0) | 0.154 |
| **Sex** | 75 |  |  |  |  |  | 0.308 |
| Female |  | 42 (56.0%) | 17 (63.0%) | 4 (40.0%) | 17 (63.0%) | 4 (36.4%) |  |
| Male |  | 33 (44.0%) | 10 (37.0%) | 6 (60.0%) | 10 (37.0%) | 7 (63.6%) |  |
| **Race** | 75 |  |  |  |  |  | 0.574 |
| Asian |  | 1 (1.3%) | 1 (3.7%) | 0 (0.0%) | 0 (0.0%) | 0 (0.0%) |  |
| Black or African American |  | 8 (10.7%) | 5 (18.5%) | 0 (0.0%) | 2 (7.4%) | 1 (9.1%) |  |
| White |  | 66 (88.0%) | 21 (77.8%) | 10 (100.0%) | 25 (92.6%) | 10 (90.9%) |  |
| **Ethnicity** | 75 |  |  |  |  |  | 1.000 |
| Not Hispanic or Latino |  | 74 (98.7%) | 26 (96.3%) | 10 (100.0%) | 27 (100.0%) | 11 (100.0%) |  |
| Hispanic or Latino |  | 1 (1.3%) | 1 (3.7%) | 0 (0.0%) | 0 (0.0%) | 0 (0.0%) |  |
| **Aβ-PET status** | 75 |  |  |  |  |  | — |
| PiB- |  | 54 (72.0%) | 27 (100.0%) | 0 (0.0%) | 27 (100.0%) | 0 (0.0%) |  |
| PiB+ |  | 21 (28.0%) | 0 (0.0%) | 10 (100.0%) | 0 (0.0%) | 11 (100.0%) |  |
| **MCI subtype** | 75 |  |  |  |  |  | 0.019 |
| Amnestic MCI |  | 59 (78.7%) | 17 (63.0%) | 10 (100.0%) | 21 (77.8%) | 11 (100.0%) |  |
| Non-amnestic MCI |  | 16 (21.3%) | 10 (37.0%) | 0 (0.0%) | 6 (22.2%) | 0 (0.0%) |  |
| **Burden score** | 75 | 2.0 (0.0, 4.0) | 2.0 (0.0, 4.0) | 0.0 (0.0, 1.0) | 3.0 (1.0, 4.1) | 1.0 (0.0, 3.0) | 0.045 |
| **eGFR (CKD-EPI 2021)** | 75 | 84.2 (70.9, 91.9) | 76.8 (65.8, 90.2) | 83.8 (70.9, 90.0) | 85.8 (75.0, 92.5) | 85.6 (70.6, 91.8) | 0.205 |
| **Serum creatinine**, mg/dL | 75 | 0.9 (0.8, 1.0) | 0.9 (0.8, 1.0) | 0.9 (0.8, 0.9) | 0.8 (0.7, 0.9) | 0.9 (0.6, 1.0) | 0.222 |
| **Number of medications** | 75 | 6.0 (4.0, 9.0) | 6.0 (4.0, 9.0) | 4.5 (2.0, 6.0) | 7.0 (5.0, 10.0) | 6.0 (4.0, 12.0) | 0.254 |
| **CDR Global score** | 75 |  |  |  |  |  | 0.594 |
| 0 |  | 24 (32.0%) | 10 (37.0%) | 2 (20.0%) | 10 (37.0%) | 2 (18.2%) |  |
| 0.5 |  | 51 (68.0%) | 17 (63.0%) | 8 (80.0%) | 17 (63.0%) | 9 (81.8%) |  |
| **CDR Sum of Boxes** | 75 | 1.0 (0.5, 1.5) | 0.5 (0.0, 1.0) | 1.3 (0.5, 2.0) | 0.5 (0.5, 1.0) | 1.0 (0.5, 1.5) | 0.087 |
| **PHQ-9 total score** | 75 | 3.0 (1.0, 5.0) | 3.0 (1.0, 5.0) | 1.5 (0.0, 5.0) | 3.0 (2.0, 8.0) | 3.0 (1.0, 3.0) | 0.164 |
| **PASE total score** | 74 | 89.8 (61.1, 133.4) | 67.6 (35.0, 129.7) | 117.3 (61.8, 140.3) | 109.6 (78.6, 184.6) | 86.4 (41.0, 126.0) | 0.151 |
| **PACC** | 73 | 0.8 (-2.3, 2.1) | 1.3 (-1.2, 3.4) | 0.0 (-4.8, 1.1) | 1.3 (0.1, 2.3) | -3.2 (-6.7, 0.1) | 0.004 |
| **Hippocampal volume**, mm<sup>3</sup> | 61 | 7,366.8 (6,630.7, 7,729.9) | 7,418.0 (6,461.3, 7,696.6) | 6,876.8 (6,472.8, 7,581.4) | 7,676.2 (7,009.8, 8,259.4) | 6,775.6 (6,339.8, 7,366.8) | 0.024 |
| **BVMT-R Delayed Recall** | 74 | 7.0 (4.0, 9.0) | 8.0 (5.0, 9.0) | 4.5 (3.0, 7.0) | 7.5 (5.0, 9.0) | 4.0 (4.0, 8.0) | 0.027 |
| **CVLT-II Long-Delay Free Recall** | 74 | 8.0 (6.0, 10.0) | 9.0 (6.0, 12.0) | 5.5 (3.0, 7.0) | 9.0 (8.0, 11.0) | 6.0 (2.0, 7.0) | 4.87 × 10<sup>-4</sup> |
| **Cortical volume**, mm<sup>3</sup> | 61 | 408,910.9 (397,107.8, 428,692.6) | 404,250.2 (389,767.0, 421,862.6) | 410,548.0 (399,772.8, 417,455.5) | 429,615.1 (408,669.5, 440,871.1) | 399,908.0 (393,800.6, 407,737.9) | 0.003 |

Restricted to the 75 participants with available Aβ-PET status. Values are median (Q1, Q3) for continuous variables and n (%) for categorical variables. N indicates the number of participants with available data. p-values are from Kruskal-Wallis rank-sum tests for continuous variables and from Pearson's χ<sup>2</sup> or Fisher's exact tests for categorical variables; p < 0.05 is shown in bold. Abbreviations: Aβ, amyloid-β; BVMT-R, Brief Visuospatial Memory Test-Revised; CDR, Clinical Dementia Rating; CKD-EPI, Chronic Kidney Disease Epidemiology Collaboration; CVLT-II, California Verbal Learning Test-Second Edition; eGFR, estimated glomerular filtration rate; MCI, mild cognitive impairment; PACC, Preclinical Alzheimer's Cognitive Composite; PASE, Physical Activity Scale for the Elderly; PET, positron emission tomography; PHQ-9, Patient Health Questionnaire-9; PiB, Pittsburgh compound B.

**Supplemental Table 3.** Age- and sex-adjusted differences in baseline plasma protein levels between lithium and placebo groups (Lithium, n = 36; Placebo, n = 37)

| Protein | Mean log2 NPQ | Coefficient (log2) | Fold change | t | p-value | FDR |
|---|---|---|---|---|---|---|
| CSF2 | 14.85 | -0.341 | 0.79 | -2.69 | **0.009** | 0.900 |
| IL10 | 12.39 | 0.560 | 1.47 | 2.37 | **0.020** | 0.900 |
| MME | 9.73 | 1.086 | 2.12 | 2.33 | **0.022** | 0.900 |
| CALB2 | 11.13 | -0.252 | 0.84 | -2.04 | **0.045** | 0.900 |
| S100B | 12.19 | -0.279 | 0.82 | -2.00 | **0.049** | 0.900 |
| CX3CL1 | 13.53 | -0.278 | 0.82 | -1.99 | 0.051 | 0.900 |
| IL17A | 11.71 | 0.713 | 1.64 | 1.97 | 0.053 | 0.900 |
| CNTN2 | 9.55 | 0.369 | 1.29 | 1.89 | 0.063 | 0.900 |
| TARDBP | 13.89 | 0.460 | 1.38 | 1.83 | 0.072 | 0.900 |
| AB40 | 11.90 | -0.247 | 0.84 | -1.79 | 0.077 | 0.900 |
| VEGFD | 13.21 | -0.810 | 0.57 | -1.75 | 0.084 | 0.900 |
| NPTX2 | 13.29 | -0.168 | 0.89 | -1.69 | 0.096 | 0.900 |
| AB38 | 10.47 | -0.251 | 0.84 | -1.62 | 0.109 | 0.900 |
| GDF15 | 12.75 | 0.435 | 1.35 | 1.61 | 0.111 | 0.900 |
| CCL17 | 13.85 | -0.458 | 0.73 | -1.59 | 0.115 | 0.900 |
| FCN2 | 12.57 | -0.208 | 0.87 | -1.56 | 0.122 | 0.900 |
| AGRN | 13.41 | -0.118 | 0.92 | -1.51 | 0.136 | 0.900 |
| TEK | 13.15 | -0.141 | 0.91 | -1.51 | 0.136 | 0.900 |
| FOLR1 | 10.42 | -0.153 | 0.90 | -1.45 | 0.151 | 0.900 |
| CCL22 | 13.20 | -0.178 | 0.88 | -1.40 | 0.165 | 0.900 |
| NPY | 15.17 | 0.629 | 1.55 | 1.38 | 0.171 | 0.900 |
| S100A12 | 12.65 | 0.331 | 1.26 | 1.37 | 0.176 | 0.900 |
| SQSTM1 | 11.51 | 0.331 | 1.26 | 1.37 | 0.176 | 0.900 |
| CST3 | 13.42 | -0.089 | 0.94 | -1.33 | 0.187 | 0.900 |
| CHIT1 | 13.28 | -1.022 | 0.49 | -1.32 | 0.190 | 0.900 |
| NRGN | 16.36 | 0.644 | 1.56 | 1.29 | 0.202 | 0.900 |
| HTT | 13.54 | 0.378 | 1.30 | 1.27 | 0.207 | 0.900 |
| PSEN1 | 12.50 | 0.259 | 1.20 | 1.25 | 0.215 | 0.900 |
| PARK7 | 15.03 | -0.634 | 0.64 | -1.19 | 0.238 | 0.900 |
| FGF2 | 14.27 | 0.328 | 1.26 | 1.17 | 0.246 | 0.900 |
| TREM1 | 11.70 | -0.120 | 0.92 | -1.16 | 0.250 | 0.900 |
| KLK6 | 12.34 | -0.138 | 0.91 | -1.13 | 0.262 | 0.900 |
| ICAM1 | 13.21 | -0.120 | 0.92 | -1.11 | 0.270 | 0.900 |
| IGF1R | 12.44 | -0.067 | 0.95 | -1.10 | 0.275 | 0.900 |
| SLIT2 | 13.38 | -0.115 | 0.92 | -1.08 | 0.282 | 0.900 |
| GFAP | 13.11 | -0.142 | 0.91 | -1.07 | 0.289 | 0.900 |
| P_SNCA_129 | 14.69 | 0.243 | 1.18 | 1.07 | 0.289 | 0.900 |
| PDLIM5 | 9.94 | -0.508 | 0.70 | -1.06 | 0.293 | 0.900 |
| P_TAU_181 | 12.77 | 0.157 | 1.11 | 1.04 | 0.302 | 0.900 |
| YWHAG | 13.00 | -0.179 | 0.88 | -1.03 | 0.305 | 0.900 |
| NEFL | 11.75 | -0.132 | 0.91 | -1.01 | 0.318 | 0.900 |
| OLIGO_SNCA | 14.23 | 0.375 | 1.30 | 0.99 | 0.323 | 0.900 |
| POSTN | 13.05 | 0.443 | 1.36 | 0.99 | 0.325 | 0.900 |
| TREM2 | 12.95 | -0.135 | 0.91 | -0.97 | 0.337 | 0.900 |
| MAPT | 11.94 | 0.224 | 1.17 | 0.96 | 0.339 | 0.900 |
| CCL11 | 13.69 | 0.124 | 1.09 | 0.96 | 0.342 | 0.900 |
| KDR | 12.43 | -0.349 | 0.79 | -0.93 | 0.354 | 0.900 |
| RUVBL2 | 11.80 | 0.115 | 1.08 | 0.92 | 0.358 | 0.900 |
| GDNF | 11.00 | 0.403 | 1.32 | 0.89 | 0.377 | 0.900 |
| CCL2 | 10.91 | -0.094 | 0.94 | -0.89 | 0.378 | 0.900 |
| VCAM1 | 13.58 | -0.072 | 0.95 | -0.87 | 0.385 | 0.900 |
| REST | 12.80 | 0.172 | 1.13 | 0.87 | 0.388 | 0.900 |
| PTN | 5.32 | 0.167 | 1.12 | 0.85 | 0.397 | 0.900 |
| IL9 | 10.39 | 0.171 | 1.13 | 0.84 | 0.401 | 0.900 |
| GOT1 | 11.75 | 0.078 | 1.06 | 0.83 | 0.408 | 0.900 |
| YWHAZ | 12.76 | -0.365 | 0.78 | -0.82 | 0.412 | 0.900 |
| BDNF | 14.51 | 0.225 | 1.17 | 0.82 | 0.413 | 0.900 |
| CD40LG | 13.48 | 0.201 | 1.15 | 0.81 | 0.421 | 0.900 |
| SNCA | 14.51 | 0.127 | 1.09 | 0.80 | 0.427 | 0.900 |
| IL4 | 11.36 | 0.233 | 1.18 | 0.80 | 0.428 | 0.900 |
| SNCB | 11.46 | 0.058 | 1.04 | 0.79 | 0.432 | 0.900 |
| IL5 | 13.18 | -0.184 | 0.88 | -0.77 | 0.443 | 0.904 |
| GDI1 | 12.31 | -0.140 | 0.91 | -0.76 | 0.449 | 0.904 |
| IGFBP7 | 13.17 | -0.054 | 0.96 | -0.74 | 0.464 | 0.910 |
| CXCL8 | 11.22 | -0.074 | 0.95 | -0.72 | 0.473 | 0.910 |
| BACE1 | 13.36 | -0.064 | 0.96 | -0.70 | 0.483 | 0.910 |
| VGF | 12.52 | -0.255 | 0.84 | -0.69 | 0.490 | 0.910 |
| MDH1 | 13.20 | 0.107 | 1.08 | 0.69 | 0.492 | 0.910 |
| MSLN | 13.64 | -0.149 | 0.90 | -0.69 | 0.494 | 0.910 |
| P_TAU_231 | 13.01 | 0.114 | 1.08 | 0.65 | 0.519 | 0.929 |
| P_TDP43_409 | 10.94 | 0.090 | 1.06 | 0.65 | 0.520 | 0.929 |
| HBA1 | 7.52 | 0.328 | 1.26 | 0.62 | 0.535 | 0.944 |
| P_TAU_217 | 11.70 | -0.066 | 0.96 | -0.60 | 0.553 | 0.948 |
| CCL3 | 12.26 | 0.149 | 1.11 | 0.59 | 0.559 | 0.948 |
| CCL26 | 11.60 | -0.076 | 0.95 | -0.59 | 0.560 | 0.948 |
| SAA1 | 12.48 | -0.220 | 0.86 | -0.56 | 0.574 | 0.960 |
| IL2 | 11.30 | 0.073 | 1.05 | 0.54 | 0.591 | 0.962 |
| TNF | 13.16 | 0.059 | 1.04 | 0.54 | 0.591 | 0.962 |
| CD63 | 12.56 | 0.078 | 1.06 | 0.51 | 0.615 | 0.962 |
| BASP1 | 12.62 | -0.154 | 0.90 | -0.48 | 0.630 | 0.962 |
| ENO2 | 13.88 | 0.114 | 1.08 | 0.47 | 0.637 | 0.962 |
| SFTPD | 13.47 | 0.083 | 1.06 | 0.43 | 0.665 | 0.962 |
| IL7 | 13.29 | 0.070 | 1.05 | 0.43 | 0.671 | 0.962 |
| VSNL1 | 12.16 | 0.036 | 1.03 | 0.42 | 0.672 | 0.962 |
| VEGFA | 13.67 | -0.046 | 0.97 | -0.42 | 0.673 | 0.962 |
| CCL13 | 13.32 | -0.059 | 0.96 | -0.42 | 0.673 | 0.962 |
| IL15 | 13.08 | -0.039 | 0.97 | -0.42 | 0.676 | 0.962 |
| SMOC1 | 12.58 | -0.145 | 0.90 | -0.41 | 0.679 | 0.962 |
| NPTX1 | 11.86 | -0.042 | 0.97 | -0.41 | 0.682 | 0.962 |
| FLT1 | 13.38 | -0.044 | 0.97 | -0.40 | 0.688 | 0.962 |
| IL18 | 13.61 | -0.091 | 0.94 | -0.40 | 0.692 | 0.962 |
| TAFA5 | 10.94 | -0.090 | 0.94 | -0.39 | 0.698 | 0.962 |
| FABP3 | 13.26 | 0.042 | 1.03 | 0.38 | 0.704 | 0.962 |
| CRH | 12.77 | 0.109 | 1.08 | 0.35 | 0.724 | 0.974 |
| PGF | 13.61 | -0.028 | 0.98 | -0.34 | 0.733 | 0.974 |
| NEFH | 18.11 | -0.183 | 0.88 | -0.34 | 0.736 | 0.974 |
| PGK1 | 11.87 | 0.111 | 1.08 | 0.33 | 0.746 | 0.974 |
| AB42 | 13.07 | 0.068 | 1.05 | 0.32 | 0.754 | 0.974 |
| TIMP3 | 14.81 | 0.085 | 1.06 | 0.28 | 0.779 | 0.974 |
| NPTXR | 11.32 | -0.033 | 0.98 | -0.28 | 0.779 | 0.974 |
| IL6R | 12.24 | 0.029 | 1.02 | 0.28 | 0.781 | 0.974 |
| SOD1 | 12.37 | 0.054 | 1.04 | 0.27 | 0.785 | 0.974 |
| ANXA5 | 14.16 | 0.102 | 1.07 | 0.25 | 0.802 | 0.974 |
| SNAP25 | 12.85 | 0.009 | 1.01 | 0.24 | 0.813 | 0.974 |
| PDGFRB | 12.96 | 0.041 | 1.03 | 0.22 | 0.827 | 0.974 |
| SFRP1 | 15.73 | 0.070 | 1.05 | 0.21 | 0.831 | 0.974 |
| DDC | 11.75 | 0.037 | 1.03 | 0.21 | 0.834 | 0.974 |
| UCHL1 | 11.95 | 0.008 | 1.01 | 0.21 | 0.836 | 0.974 |
| IL12P70 | 13.68 | 0.053 | 1.04 | 0.21 | 0.836 | 0.974 |
| CRP | 12.22 | -0.061 | 0.96 | -0.18 | 0.854 | 0.979 |
| CHI3L1 | 13.44 | -0.045 | 0.97 | -0.17 | 0.865 | 0.979 |
| IFNG | 12.51 | -0.042 | 0.97 | -0.17 | 0.866 | 0.979 |
| IL1B | 11.12 | -0.014 | 0.99 | -0.16 | 0.876 | 0.979 |
| CXCL10 | 11.33 | 0.022 | 1.02 | 0.15 | 0.878 | 0.979 |
| NGF | 8.83 | -0.018 | 0.99 | -0.12 | 0.907 | 0.989 |
| CCL4 | 12.35 | 0.029 | 1.02 | 0.11 | 0.912 | 0.989 |
| PRDX6 | 12.70 | -0.030 | 0.98 | -0.10 | 0.924 | 0.989 |
| IL16 | 12.99 | -0.010 | 0.99 | -0.09 | 0.928 | 0.989 |
| APOE4 | 4.92 | 0.122 | 1.09 | 0.09 | 0.931 | 0.989 |
| ARSA | 12.65 | -0.023 | 0.98 | -0.08 | 0.937 | 0.989 |
| IL13 | 13.00 | -0.029 | 0.98 | -0.07 | 0.942 | 0.989 |
| CXCL1 | 12.12 | 0.014 | 1.01 | 0.06 | 0.954 | 0.993 |
| IL33 | 12.53 | -0.004 | 1.00 | -0.04 | 0.969 | 0.999 |
| APOE | 13.34 | -0.003 | 1.00 | -0.01 | 0.990 | 0.999 |
| ACHE | 12.96 | 0.000 | 1.00 | -0.00 | 0.997 | 0.999 |
| IL6 | 12.91 | -0.001 | 1.00 | -0.00 | 0.998 | 0.999 |
| UBB | 12.89 | 0.000 | 1.00 | 0.00 | 0.999 | 0.999 |

Estimates are from linear models of baseline log2 NPQ adjusted for age and sex, fitted with limma with empirical Bayes moderation (trend and robust). The coefficient is the log2 difference (lithium minus placebo); fold change = 2^coefficient. FDR, Benjamini-Hochberg adjustment across all 127 proteins; p < 0.05 shown in bold. Proteins are ordered by p-value and use NULISA target names. These are the statistics plotted in the Figure 1a volcano plot. Abbreviations: FDR, false discovery rate; NPQ, NULISA Protein Quantification.

**Supplemental Table 4.**
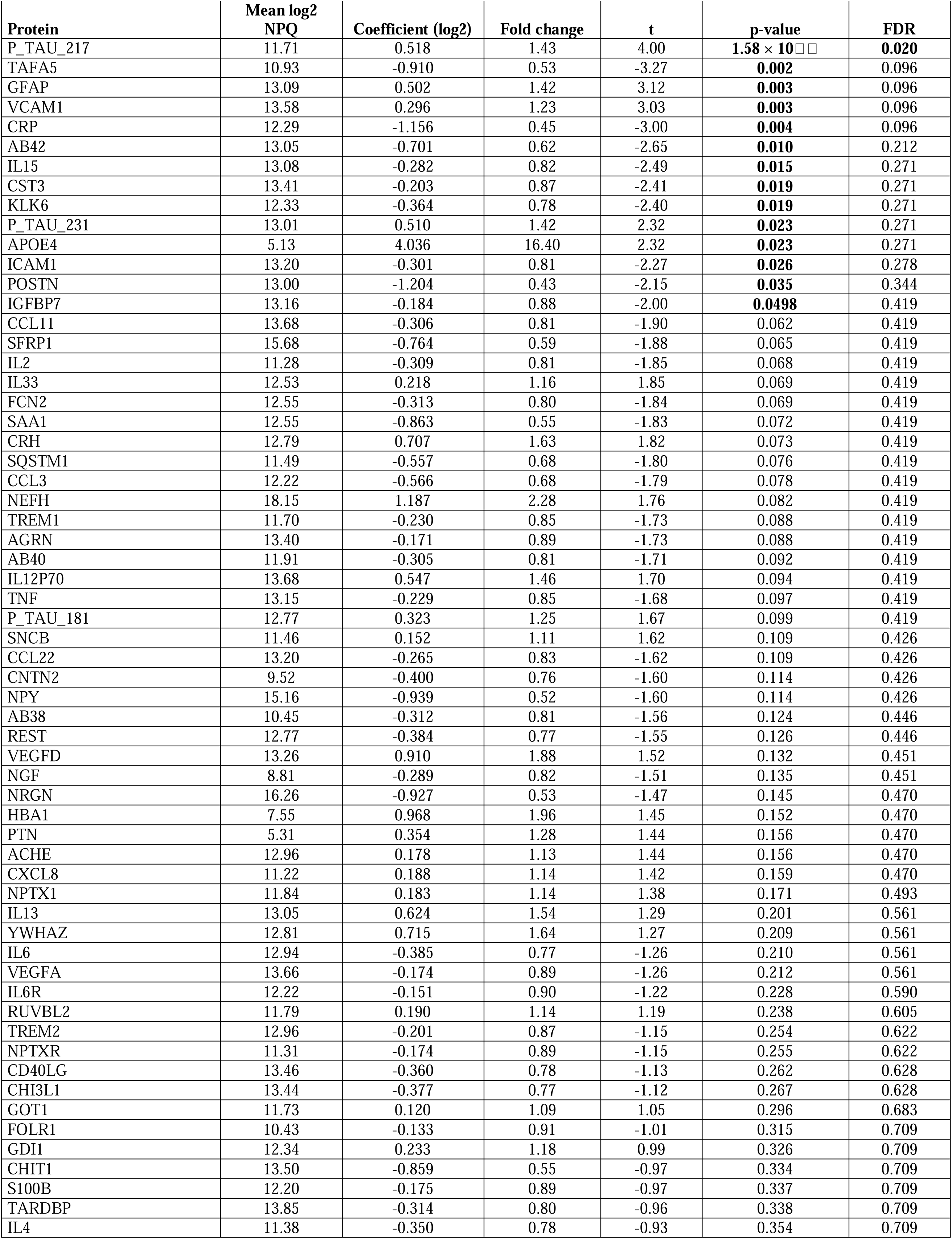
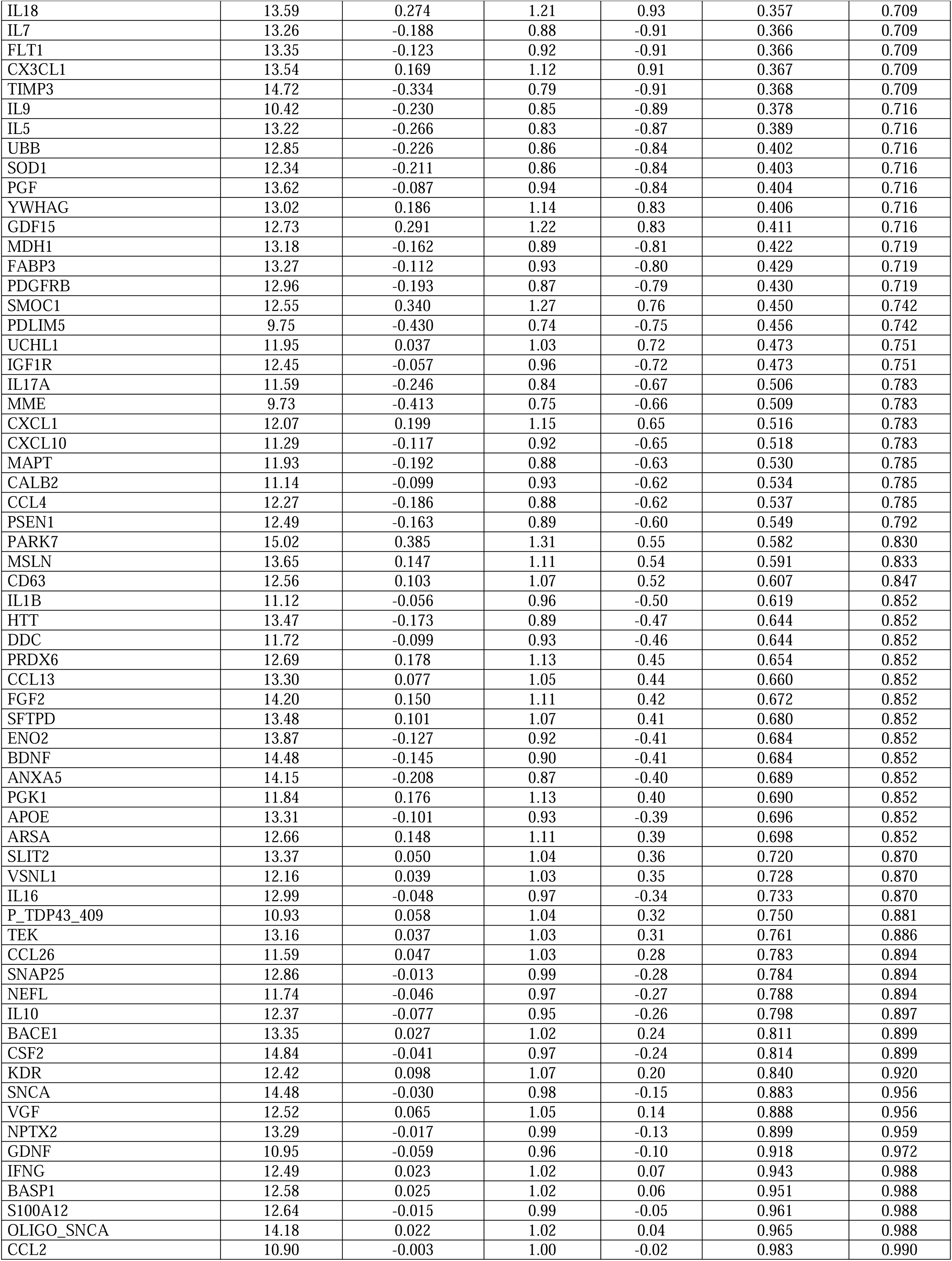
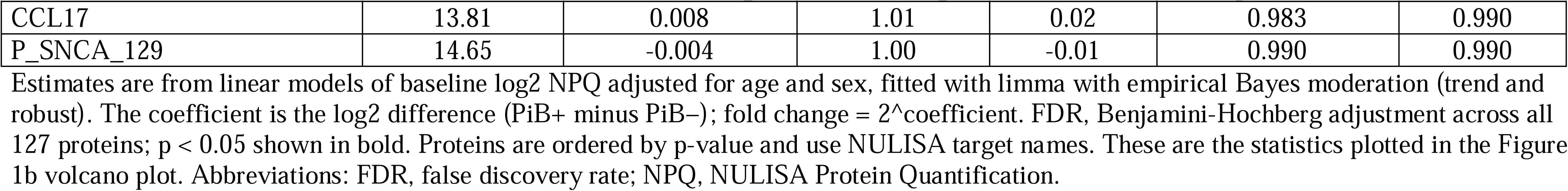
Age- and sex-adjusted differences in baseline plasma protein levels between Aβ-PET-positive and Aβ-PET-negative participants (PiB+, n = 20; PiB−, n = 50)

| **Protein** | **Mean log2 NPQ** | **Coefficient (log2)** | **Fold change** | **t** | **p-value** | **FDR** |
|---|---|---|---|---|---|---|
| P_TAU_217 | 11.71 | 0.518 | 1.43 | 4.00 | **1.58 × 10[illegible]** | **0.020** |
| TAFA5 | 10.93 | -0.910 | 0.53 | -3.27 | **0.002** | 0.096 |
| GFAP | 13.09 | 0.502 | 1.42 | 3.12 | **0.003** | 0.096 |
| VCAM1 | 13.58 | 0.296 | 1.23 | 3.03 | **0.003** | 0.096 |
| CRP | 12.29 | -1.156 | 0.45 | -3.00 | **0.004** | 0.096 |
| AB42 | 13.05 | -0.701 | 0.62 | -2.65 | **0.010** | 0.212 |
| IL15 | 13.08 | -0.282 | 0.82 | -2.49 | **0.015** | 0.271 |
| CST3 | 13.41 | -0.203 | 0.87 | -2.41 | **0.019** | 0.271 |
| KLK6 | 12.33 | -0.364 | 0.78 | -2.40 | **0.019** | 0.271 |
| P_TAU_231 | 13.01 | 0.510 | 1.42 | 2.32 | **0.023** | 0.271 |
| APOE4 | 5.13 | 4.036 | 16.40 | 2.32 | **0.023** | 0.271 |
| ICAM1 | 13.20 | -0.301 | 0.81 | -2.27 | **0.026** | 0.278 |
| POSTN | 13.00 | -1.204 | 0.43 | -2.15 | **0.035** | 0.344 |
| IGFBP7 | 13.16 | -0.184 | 0.88 | -2.00 | **0.0498** | 0.419 |
| CCL11 | 13.68 | -0.306 | 0.81 | -1.90 | 0.062 | 0.419 |
| SFRP1 | 15.68 | -0.764 | 0.59 | -1.88 | 0.065 | 0.419 |
| IL2 | 11.28 | -0.309 | 0.81 | -1.85 | 0.068 | 0.419 |
| IL33 | 12.53 | 0.218 | 1.16 | 1.85 | 0.069 | 0.419 |
| FCN2 | 12.55 | -0.313 | 0.80 | -1.84 | 0.069 | 0.419 |
| SAA1 | 12.55 | -0.863 | 0.55 | -1.83 | 0.072 | 0.419 |
| CRH | 12.79 | 0.707 | 1.63 | 1.82 | 0.073 | 0.419 |
| SQSTM1 | 11.49 | -0.557 | 0.68 | -1.80 | 0.076 | 0.419 |
| CCL3 | 12.22 | -0.566 | 0.68 | -1.79 | 0.078 | 0.419 |
| NEFH | 18.15 | 1.187 | 2.28 | 1.76 | 0.082 | 0.419 |
| TREM1 | 11.70 | -0.230 | 0.85 | -1.73 | 0.088 | 0.419 |
| AGRN | 13.40 | -0.171 | 0.89 | -1.73 | 0.088 | 0.419 |
| AB40 | 11.91 | -0.305 | 0.81 | -1.71 | 0.092 | 0.419 |
| IL12P70 | 13.68 | 0.547 | 1.46 | 1.70 | 0.094 | 0.419 |
| TNF | 13.15 | -0.229 | 0.85 | -1.68 | 0.097 | 0.419 |
| P_TAU_181 | 12.77 | 0.323 | 1.25 | 1.67 | 0.099 | 0.419 |
| SNCB | 11.46 | 0.152 | 1.11 | 1.62 | 0.109 | 0.426 |
| CCL22 | 13.20 | -0.265 | 0.83 | -1.62 | 0.109 | 0.426 |
| CNTN2 | 9.52 | -0.400 | 0.76 | -1.60 | 0.114 | 0.426 |
| NPY | 15.16 | -0.939 | 0.52 | -1.60 | 0.114 | 0.426 |
| AB38 | 10.45 | -0.312 | 0.81 | -1.56 | 0.124 | 0.446 |
| REST | 12.77 | -0.384 | 0.77 | -1.55 | 0.126 | 0.446 |
| VEGFD | 13.26 | 0.910 | 1.88 | 1.52 | 0.132 | 0.451 |
| NGF | 8.81 | -0.289 | 0.82 | -1.51 | 0.135 | 0.451 |
| NRGN | 16.26 | -0.927 | 0.53 | -1.47 | 0.145 | 0.470 |
| HBA1 | 7.55 | 0.968 | 1.96 | 1.45 | 0.152 | 0.470 |
| PTN | 5.31 | 0.354 | 1.28 | 1.44 | 0.156 | 0.470 |
| ACHE | 12.96 | 0.178 | 1.13 | 1.44 | 0.156 | 0.470 |
| CXCL8 | 11.22 | 0.188 | 1.14 | 1.42 | 0.159 | 0.470 |
| NPTX1 | 11.84 | 0.183 | 1.14 | 1.38 | 0.171 | 0.493 |
| IL13 | 13.05 | 0.624 | 1.54 | 1.29 | 0.201 | 0.561 |
| YWHAZ | 12.81 | 0.715 | 1.64 | 1.27 | 0.209 | 0.561 |
| IL6 | 12.94 | -0.385 | 0.77 | -1.26 | 0.210 | 0.561 |
| VEGFA | 13.66 | -0.174 | 0.89 | -1.26 | 0.212 | 0.561 |
| IL6R | 12.22 | -0.151 | 0.90 | -1.22 | 0.228 | 0.590 |
| RUVBL2 | 11.79 | 0.190 | 1.14 | 1.19 | 0.238 | 0.605 |
| TREM2 | 12.96 | -0.201 | 0.87 | -1.15 | 0.254 | 0.622 |
| NPTXR | 11.31 | -0.174 | 0.89 | -1.15 | 0.255 | 0.622 |
| CD40LG | 13.46 | -0.360 | 0.78 | -1.13 | 0.262 | 0.628 |
| CHI3L1 | 13.44 | -0.377 | 0.77 | -1.12 | 0.267 | 0.628 |
| GOT1 | 11.73 | 0.120 | 1.09 | 1.05 | 0.296 | 0.683 |
| FOLR1 | 10.43 | -0.133 | 0.91 | -1.01 | 0.315 | 0.709 |
| GDI1 | 12.34 | 0.233 | 1.18 | 0.99 | 0.326 | 0.709 |
| CHIT1 | 13.50 | -0.859 | 0.55 | -0.97 | 0.334 | 0.709 |
| S100B | 12.20 | -0.175 | 0.89 | -0.97 | 0.337 | 0.709 |
| TARDBP | 13.85 | -0.314 | 0.80 | -0.96 | 0.338 | 0.709 |
| IL4 | 11.38 | -0.350 | 0.78 | -0.93 | 0.354 | 0.709 |
| IL18 | 13.59 | 0.274 | 1.21 | 0.93 | 0.357 | 0.709 |
| IL7 | 13.26 | -0.188 | 0.88 | -0.91 | 0.366 | 0.709 |
| FLT1 | 13.35 | -0.123 | 0.92 | -0.91 | 0.366 | 0.709 |
| CX3CL1 | 13.54 | 0.169 | 1.12 | 0.91 | 0.367 | 0.709 |
| TIMP3 | 14.72 | -0.334 | 0.79 | -0.91 | 0.368 | 0.709 |
| IL9 | 10.42 | -0.230 | 0.85 | -0.89 | 0.378 | 0.716 |
| IL5 | 13.22 | -0.266 | 0.83 | -0.87 | 0.389 | 0.716 |
| UBB | 12.85 | -0.226 | 0.86 | -0.84 | 0.402 | 0.716 |
| SOD1 | 12.34 | -0.211 | 0.86 | -0.84 | 0.403 | 0.716 |
| PGF | 13.62 | -0.087 | 0.94 | -0.84 | 0.404 | 0.716 |
| YWHAG | 13.02 | 0.186 | 1.14 | 0.83 | 0.406 | 0.716 |
| GDF15 | 12.73 | 0.291 | 1.22 | 0.83 | 0.411 | 0.716 |
| MDH1 | 13.18 | -0.162 | 0.89 | -0.81 | 0.422 | 0.719 |
| FABP3 | 13.27 | -0.112 | 0.93 | -0.80 | 0.429 | 0.719 |
| PDGFRB | 12.96 | -0.193 | 0.87 | -0.79 | 0.430 | 0.719 |
| SMOC1 | 12.55 | 0.340 | 1.27 | 0.76 | 0.450 | 0.742 |
| PDLIM5 | 9.75 | -0.430 | 0.74 | -0.75 | 0.456 | 0.742 |
| UCHL1 | 11.95 | 0.037 | 1.03 | 0.72 | 0.473 | 0.751 |
| IGF1R | 12.45 | -0.057 | 0.96 | -0.72 | 0.473 | 0.751 |
| IL17A | 11.59 | -0.246 | 0.84 | -0.67 | 0.506 | 0.783 |
| MME | 9.73 | -0.413 | 0.75 | -0.66 | 0.509 | 0.783 |
| CXCL1 | 12.07 | 0.199 | 1.15 | 0.65 | 0.516 | 0.783 |
| CXCL10 | 11.29 | -0.117 | 0.92 | -0.65 | 0.518 | 0.783 |
| MAPT | 11.93 | -0.192 | 0.88 | -0.63 | 0.530 | 0.785 |
| CALB2 | 11.14 | -0.099 | 0.93 | -0.62 | 0.534 | 0.785 |
| CCL4 | 12.27 | -0.186 | 0.88 | -0.62 | 0.537 | 0.785 |
| PSEN1 | 12.49 | -0.163 | 0.89 | -0.60 | 0.549 | 0.792 |
| PARK7 | 15.02 | 0.385 | 1.31 | 0.55 | 0.582 | 0.830 |
| MSLN | 13.65 | 0.147 | 1.11 | 0.54 | 0.591 | 0.833 |
| CD63 | 12.56 | 0.103 | 1.07 | 0.52 | 0.607 | 0.847 |
| IL1B | 11.12 | -0.056 | 0.96 | -0.50 | 0.619 | 0.852 |
| HTT | 13.47 | -0.173 | 0.89 | -0.47 | 0.644 | 0.852 |
| DDC | 11.72 | -0.099 | 0.93 | -0.46 | 0.644 | 0.852 |
| PRDX6 | 12.69 | 0.178 | 1.13 | 0.45 | 0.654 | 0.852 |
| CCL13 | 13.30 | 0.077 | 1.05 | 0.44 | 0.660 | 0.852 |
| FGF2 | 14.20 | 0.150 | 1.11 | 0.42 | 0.672 | 0.852 |
| SFTPD | 13.48 | 0.101 | 1.07 | 0.41 | 0.680 | 0.852 |
| ENO2 | 13.87 | -0.127 | 0.92 | -0.41 | 0.684 | 0.852 |
| BDNF | 14.48 | -0.145 | 0.90 | -0.41 | 0.684 | 0.852 |
| ANXA5 | 14.15 | -0.208 | 0.87 | -0.40 | 0.689 | 0.852 |
| PGK1 | 11.84 | 0.176 | 1.13 | 0.40 | 0.690 | 0.852 |
| APOE | 13.31 | -0.101 | 0.93 | -0.39 | 0.696 | 0.852 |
| ARSA | 12.66 | 0.148 | 1.11 | 0.39 | 0.698 | 0.852 |
| SLIT2 | 13.37 | 0.050 | 1.04 | 0.36 | 0.720 | 0.870 |
| VSNL1 | 12.16 | 0.039 | 1.03 | 0.35 | 0.728 | 0.870 |
| IL16 | 12.99 | -0.048 | 0.97 | -0.34 | 0.733 | 0.870 |
| P_TDP43_409 | 10.93 | 0.058 | 1.04 | 0.32 | 0.750 | 0.881 |
| TEK | 13.16 | 0.037 | 1.03 | 0.31 | 0.761 | 0.886 |
| CCL26 | 11.59 | 0.047 | 1.03 | 0.28 | 0.783 | 0.894 |
| SNAP25 | 12.86 | -0.013 | 0.99 | -0.28 | 0.784 | 0.894 |
| NEFL | 11.74 | -0.046 | 0.97 | -0.27 | 0.788 | 0.894 |
| IL10 | 12.37 | -0.077 | 0.95 | -0.26 | 0.798 | 0.897 |
| BACE1 | 13.35 | 0.027 | 1.02 | 0.24 | 0.811 | 0.899 |
| CSF2 | 14.84 | -0.041 | 0.97 | -0.24 | 0.814 | 0.899 |
| KDR | 12.42 | 0.098 | 1.07 | 0.20 | 0.840 | 0.920 |
| SNCA | 14.48 | -0.030 | 0.98 | -0.15 | 0.883 | 0.956 |
| VGF | 12.52 | 0.065 | 1.05 | 0.14 | 0.888 | 0.956 |
| NPTX2 | 13.29 | -0.017 | 0.99 | -0.13 | 0.899 | 0.959 |
| GDNF | 10.95 | -0.059 | 0.96 | -0.10 | 0.918 | 0.972 |
| IFNG | 12.49 | 0.023 | 1.02 | 0.07 | 0.943 | 0.988 |
| BASP1 | 12.58 | 0.025 | 1.02 | 0.06 | 0.951 | 0.988 |
| S100A12 | 12.64 | -0.015 | 0.99 | -0.05 | 0.961 | 0.988 |
| OLIGO_SNCA | 14.18 | 0.022 | 1.02 | 0.04 | 0.965 | 0.988 |
| CCL2 | 10.90 | -0.003 | 1.00 | -0.02 | 0.983 | 0.990 |
| CCL17 | 13.81 | 0.008 | 1.01 | 0.02 | 0.983 | 0.990 |
| P_SNCA_129 | 14.65 | -0.004 | 1.00 | -0.01 | 0.990 | 0.990 |

Estimates are from linear models of baseline log2 NPQ adjusted for age and sex, fitted with limma with empirical Bayes moderation (trend and robust). The coefficient is the log2 difference (PiB+ minus PiB−); fold change = 2^coefficient. FDR, Benjamini-Hochberg adjustment across all 127 proteins; p < 0.05 shown in bold. Proteins are ordered by p-value and use NULISA target names. These are the statistics plotted in the Figure 1b volcano plot. Abbreviations: FDR, false discovery rate; NPQ, NULISA Protein Quantification.

**Supplemental Table 5.**
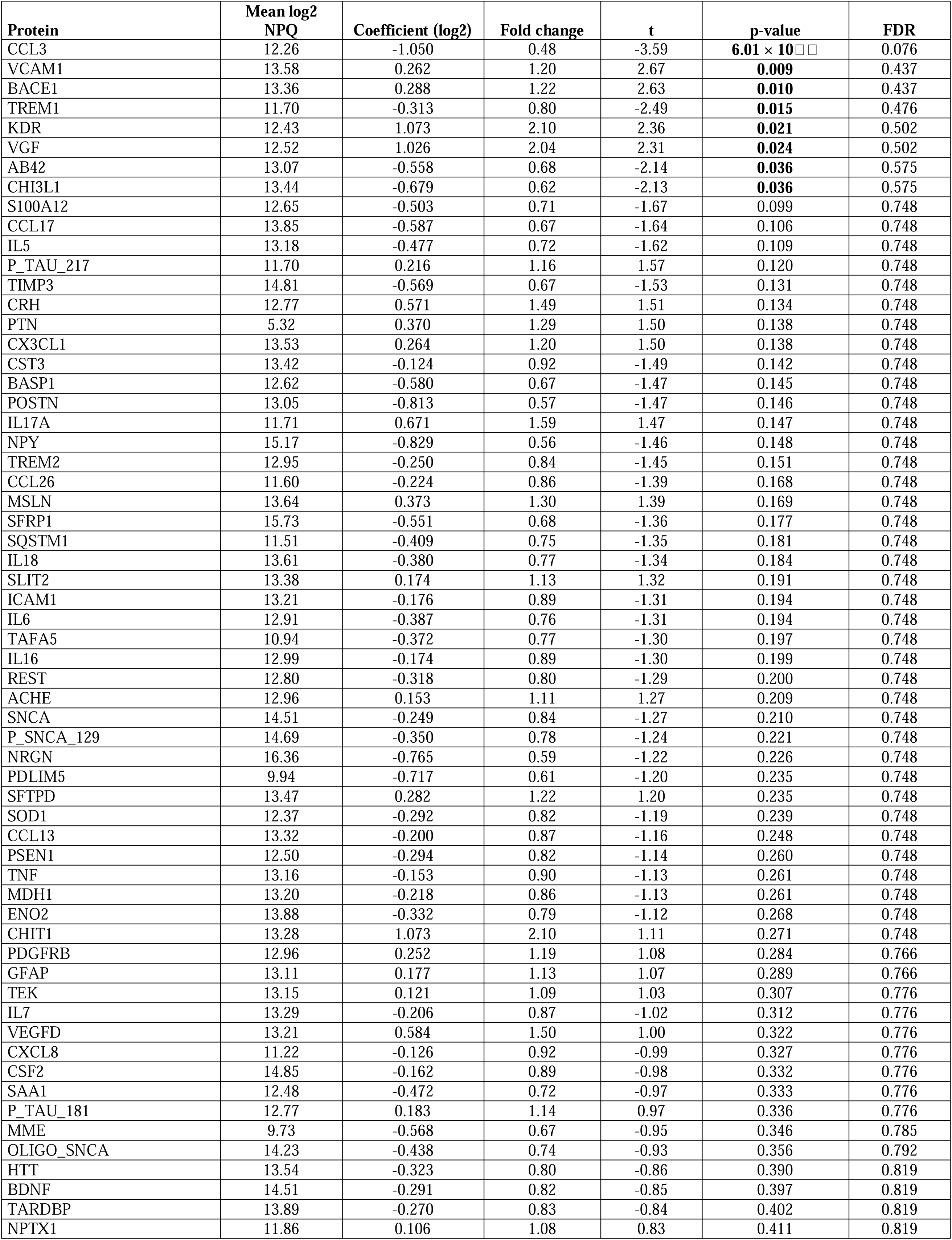
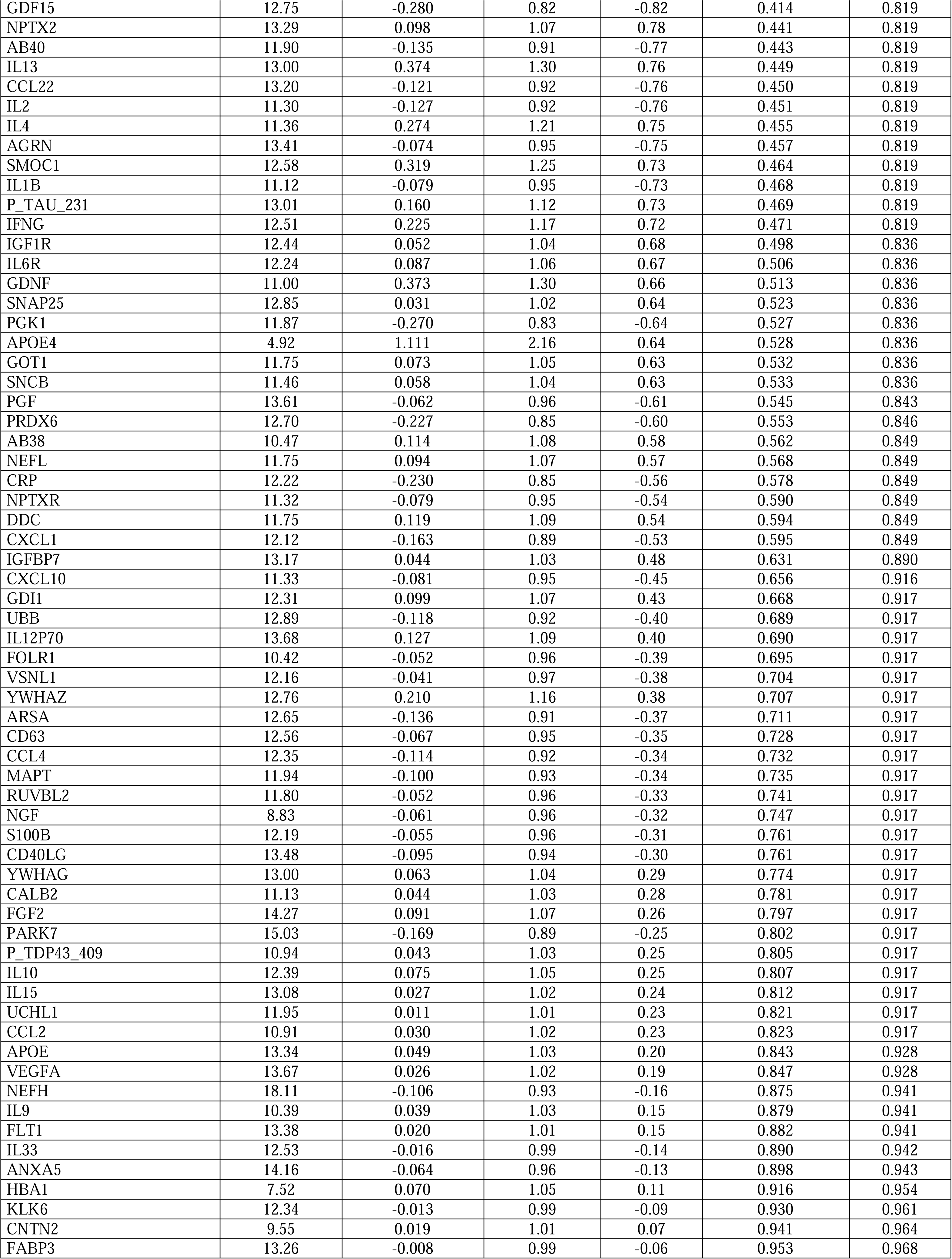
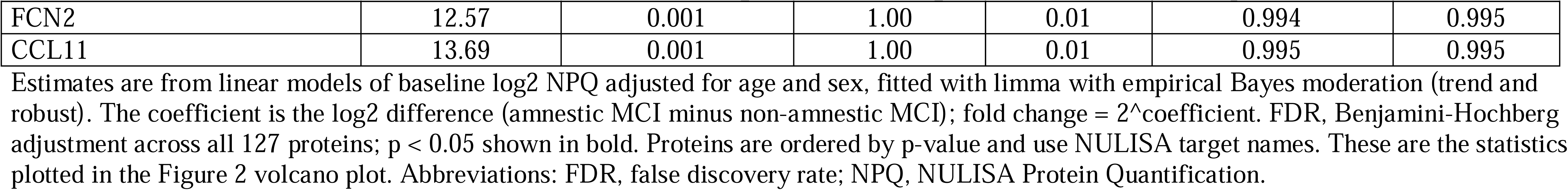
Age- and sex-adjusted differences in baseline plasma protein levels between amnestic and non-amnestic MCI (aMCI, n = 57; non-aMCI, n = 16)

| Protein | Mean log2 NPQ | Coefficient (log2) | Fold change | t | p-value | FDR |
|---|---|---|---|---|---|---|
| CCL3 | 12.26 | -1.050 | 0.48 | -3.59 | **6.01 × 10[illegible]** | 0.076 |
| VCAM1 | 13.58 | 0.262 | 1.20 | 2.67 | **0.009** | 0.437 |
| BACE1 | 13.36 | 0.288 | 1.22 | 2.63 | **0.010** | 0.437 |
| TREM1 | 11.70 | -0.313 | 0.80 | -2.49 | **0.015** | 0.476 |
| KDR | 12.43 | 1.073 | 2.10 | 2.36 | **0.021** | 0.502 |
| VGF | 12.52 | 1.026 | 2.04 | 2.31 | **0.024** | 0.502 |
| AB42 | 13.07 | -0.558 | 0.68 | -2.14 | **0.036** | 0.575 |
| CHI3L1 | 13.44 | -0.679 | 0.62 | -2.13 | **0.036** | 0.575 |
| S100A12 | 12.65 | -0.503 | 0.71 | -1.67 | 0.099 | 0.748 |
| CCL17 | 13.85 | -0.587 | 0.67 | -1.64 | 0.106 | 0.748 |
| IL5 | 13.18 | -0.477 | 0.72 | -1.62 | 0.109 | 0.748 |
| P_TAU_217 | 11.70 | 0.216 | 1.16 | 1.57 | 0.120 | 0.748 |
| TIMP3 | 14.81 | -0.569 | 0.67 | -1.53 | 0.131 | 0.748 |
| CRH | 12.77 | 0.571 | 1.49 | 1.51 | 0.134 | 0.748 |
| PTN | 5.32 | 0.370 | 1.29 | 1.50 | 0.138 | 0.748 |
| CX3CL1 | 13.53 | 0.264 | 1.20 | 1.50 | 0.138 | 0.748 |
| CST3 | 13.42 | -0.124 | 0.92 | -1.49 | 0.142 | 0.748 |
| BASP1 | 12.62 | -0.580 | 0.67 | -1.47 | 0.145 | 0.748 |
| POSTN | 13.05 | -0.813 | 0.57 | -1.47 | 0.146 | 0.748 |
| IL17A | 11.71 | 0.671 | 1.59 | 1.47 | 0.147 | 0.748 |
| NPY | 15.17 | -0.829 | 0.56 | -1.46 | 0.148 | 0.748 |
| TREM2 | 12.95 | -0.250 | 0.84 | -1.45 | 0.151 | 0.748 |
| CCL26 | 11.60 | -0.224 | 0.86 | -1.39 | 0.168 | 0.748 |
| MSLN | 13.64 | 0.373 | 1.30 | 1.39 | 0.169 | 0.748 |
| SFRP1 | 15.73 | -0.551 | 0.68 | -1.36 | 0.177 | 0.748 |
| SQSTM1 | 11.51 | -0.409 | 0.75 | -1.35 | 0.181 | 0.748 |
| IL18 | 13.61 | -0.380 | 0.77 | -1.34 | 0.184 | 0.748 |
| SLIT2 | 13.38 | 0.174 | 1.13 | 1.32 | 0.191 | 0.748 |
| ICAM1 | 13.21 | -0.176 | 0.89 | -1.31 | 0.194 | 0.748 |
| IL6 | 12.91 | -0.387 | 0.76 | -1.31 | 0.194 | 0.748 |
| TAFA5 | 10.94 | -0.372 | 0.77 | -1.30 | 0.197 | 0.748 |
| IL16 | 12.99 | -0.174 | 0.89 | -1.30 | 0.199 | 0.748 |
| REST | 12.80 | -0.318 | 0.80 | -1.29 | 0.200 | 0.748 |
| ACHE | 12.96 | 0.153 | 1.11 | 1.27 | 0.209 | 0.748 |
| SNCA | 14.51 | -0.249 | 0.84 | -1.27 | 0.210 | 0.748 |
| P_SNCA_129 | 14.69 | -0.350 | 0.78 | -1.24 | 0.221 | 0.748 |
| NRGN | 16.36 | -0.765 | 0.59 | -1.22 | 0.226 | 0.748 |
| PDLIM5 | 9.94 | -0.717 | 0.61 | -1.20 | 0.235 | 0.748 |
| SFTPD | 13.47 | 0.282 | 1.22 | 1.20 | 0.235 | 0.748 |
| SOD1 | 12.37 | -0.292 | 0.82 | -1.19 | 0.239 | 0.748 |
| CCL13 | 13.32 | -0.200 | 0.87 | -1.16 | 0.248 | 0.748 |
| PSEN1 | 12.50 | -0.294 | 0.82 | -1.14 | 0.260 | 0.748 |
| TNF | 13.16 | -0.153 | 0.90 | -1.13 | 0.261 | 0.748 |
| MDH1 | 13.20 | -0.218 | 0.86 | -1.13 | 0.261 | 0.748 |
| ENO2 | 13.88 | -0.332 | 0.79 | -1.12 | 0.268 | 0.748 |
| CHIT1 | 13.28 | 1.073 | 2.10 | 1.11 | 0.271 | 0.748 |
| PDGFRB | 12.96 | 0.252 | 1.19 | 1.08 | 0.284 | 0.766 |
| GFAP | 13.11 | 0.177 | 1.13 | 1.07 | 0.289 | 0.766 |
| TEK | 13.15 | 0.121 | 1.09 | 1.03 | 0.307 | 0.776 |
| IL7 | 13.29 | -0.206 | 0.87 | -1.02 | 0.312 | 0.776 |
| VEGFD | 13.21 | 0.584 | 1.50 | 1.00 | 0.322 | 0.776 |
| CXCL8 | 11.22 | -0.126 | 0.92 | -0.99 | 0.327 | 0.776 |
| CSF2 | 14.85 | -0.162 | 0.89 | -0.98 | 0.332 | 0.776 |
| SAA1 | 12.48 | -0.472 | 0.72 | -0.97 | 0.333 | 0.776 |
| P_TAU_181 | 12.77 | 0.183 | 1.14 | 0.97 | 0.336 | 0.776 |
| MME | 9.73 | -0.568 | 0.67 | -0.95 | 0.346 | 0.785 |
| OLIGO_SNCA | 14.23 | -0.438 | 0.74 | -0.93 | 0.356 | 0.792 |
| HTT | 13.54 | -0.323 | 0.80 | -0.86 | 0.390 | 0.819 |
| BDNF | 14.51 | -0.291 | 0.82 | -0.85 | 0.397 | 0.819 |
| TARDBP | 13.89 | -0.270 | 0.83 | -0.84 | 0.402 | 0.819 |
| NPTX1 | 11.86 | 0.106 | 1.08 | 0.83 | 0.411 | 0.819 |
| GDF15 | 12.75 | -0.280 | 0.82 | -0.82 | 0.414 | 0.819 |
| NPTX2 | 13.29 | 0.098 | 1.07 | 0.78 | 0.441 | 0.819 |
| AB40 | 11.90 | -0.135 | 0.91 | -0.77 | 0.443 | 0.819 |
| IL13 | 13.00 | 0.374 | 1.30 | 0.76 | 0.449 | 0.819 |
| CCL22 | 13.20 | -0.121 | 0.92 | -0.76 | 0.450 | 0.819 |
| IL2 | 11.30 | -0.127 | 0.92 | -0.76 | 0.451 | 0.819 |
| IL4 | 11.36 | 0.274 | 1.21 | 0.75 | 0.455 | 0.819 |
| AGRN | 13.41 | -0.074 | 0.95 | -0.75 | 0.457 | 0.819 |
| SMOC1 | 12.58 | 0.319 | 1.25 | 0.73 | 0.464 | 0.819 |
| IL1B | 11.12 | -0.079 | 0.95 | -0.73 | 0.468 | 0.819 |
| P_TAU_231 | 13.01 | 0.160 | 1.12 | 0.73 | 0.469 | 0.819 |
| IFNG | 12.51 | 0.225 | 1.17 | 0.72 | 0.471 | 0.819 |
| IGF1R | 12.44 | 0.052 | 1.04 | 0.68 | 0.498 | 0.836 |
| IL6R | 12.24 | 0.087 | 1.06 | 0.67 | 0.506 | 0.836 |
| GDNF | 11.00 | 0.373 | 1.30 | 0.66 | 0.513 | 0.836 |
| SNAP25 | 12.85 | 0.031 | 1.02 | 0.64 | 0.523 | 0.836 |
| PGK1 | 11.87 | -0.270 | 0.83 | -0.64 | 0.527 | 0.836 |
| APOE4 | 4.92 | 1.111 | 2.16 | 0.64 | 0.528 | 0.836 |
| GOT1 | 11.75 | 0.073 | 1.05 | 0.63 | 0.532 | 0.836 |
| SNCB | 11.46 | 0.058 | 1.04 | 0.63 | 0.533 | 0.836 |
| PGF | 13.61 | -0.062 | 0.96 | -0.61 | 0.545 | 0.843 |
| PRDX6 | 12.70 | -0.227 | 0.85 | -0.60 | 0.553 | 0.846 |
| AB38 | 10.47 | 0.114 | 1.08 | 0.58 | 0.562 | 0.849 |
| NEFL | 11.75 | 0.094 | 1.07 | 0.57 | 0.568 | 0.849 |
| CRP | 12.22 | -0.230 | 0.85 | -0.56 | 0.578 | 0.849 |
| NPTXR | 11.32 | -0.079 | 0.95 | -0.54 | 0.590 | 0.849 |
| DDC | 11.75 | 0.119 | 1.09 | 0.54 | 0.594 | 0.849 |
| CXCL1 | 12.12 | -0.163 | 0.89 | -0.53 | 0.595 | 0.849 |
| IGFBP7 | 13.17 | 0.044 | 1.03 | 0.48 | 0.631 | 0.890 |
| CXCL10 | 11.33 | -0.081 | 0.95 | -0.45 | 0.656 | 0.916 |
| GDI1 | 12.31 | 0.099 | 1.07 | 0.43 | 0.668 | 0.917 |
| UBB | 12.89 | -0.118 | 0.92 | -0.40 | 0.689 | 0.917 |
| IL12P70 | 13.68 | 0.127 | 1.09 | 0.40 | 0.690 | 0.917 |
| FOLR1 | 10.42 | -0.052 | 0.96 | -0.39 | 0.695 | 0.917 |
| VSNL1 | 12.16 | -0.041 | 0.97 | -0.38 | 0.704 | 0.917 |
| YWHAZ | 12.76 | 0.210 | 1.16 | 0.38 | 0.707 | 0.917 |
| ARSA | 12.65 | -0.136 | 0.91 | -0.37 | 0.711 | 0.917 |
| CD63 | 12.56 | -0.067 | 0.95 | -0.35 | 0.728 | 0.917 |
| CCL4 | 12.35 | -0.114 | 0.92 | -0.34 | 0.732 | 0.917 |
| MAPT | 11.94 | -0.100 | 0.93 | -0.34 | 0.735 | 0.917 |
| RUVBL2 | 11.80 | -0.052 | 0.96 | -0.33 | 0.741 | 0.917 |
| NGF | 8.83 | -0.061 | 0.96 | -0.32 | 0.747 | 0.917 |
| S100B | 12.19 | -0.055 | 0.96 | -0.31 | 0.761 | 0.917 |
| CD40LG | 13.48 | -0.095 | 0.94 | -0.30 | 0.761 | 0.917 |
| YWHAG | 13.00 | 0.063 | 1.04 | 0.29 | 0.774 | 0.917 |
| CALB2 | 11.13 | 0.044 | 1.03 | 0.28 | 0.781 | 0.917 |
| FGF2 | 14.27 | 0.091 | 1.07 | 0.26 | 0.797 | 0.917 |
| PARK7 | 15.03 | -0.169 | 0.89 | -0.25 | 0.802 | 0.917 |
| P_TDP43_409 | 10.94 | 0.043 | 1.03 | 0.25 | 0.805 | 0.917 |
| IL10 | 12.39 | 0.075 | 1.05 | 0.25 | 0.807 | 0.917 |
| IL15 | 13.08 | 0.027 | 1.02 | 0.24 | 0.812 | 0.917 |
| UCHL1 | 11.95 | 0.011 | 1.01 | 0.23 | 0.821 | 0.917 |
| CCL2 | 10.91 | 0.030 | 1.02 | 0.23 | 0.823 | 0.917 |
| APOE | 13.34 | 0.049 | 1.03 | 0.20 | 0.843 | 0.928 |
| VEGFA | 13.67 | 0.026 | 1.02 | 0.19 | 0.847 | 0.928 |
| NEFH | 18.11 | -0.106 | 0.93 | -0.16 | 0.875 | 0.941 |
| IL9 | 10.39 | 0.039 | 1.03 | 0.15 | 0.879 | 0.941 |
| FLT1 | 13.38 | 0.020 | 1.01 | 0.15 | 0.882 | 0.941 |
| IL33 | 12.53 | -0.016 | 0.99 | -0.14 | 0.890 | 0.942 |
| ANXA5 | 14.16 | -0.064 | 0.96 | -0.13 | 0.898 | 0.943 |
| HBA1 | 7.52 | 0.070 | 1.05 | 0.11 | 0.916 | 0.954 |
| KLK6 | 12.34 | -0.013 | 0.99 | -0.09 | 0.930 | 0.961 |
| CNTN2 | 9.55 | 0.019 | 1.01 | 0.07 | 0.941 | 0.964 |
| FABP3 | 13.26 | -0.008 | 0.99 | -0.06 | 0.953 | 0.968 |
| FCN2 | 12.57 | 0.001 | 1.00 | 0.01 | 0.994 | 0.995 |
| CCL11 | 13.69 | 0.001 | 1.00 | 0.01 | 0.995 | 0.995 |

Estimates are from linear models of baseline log2 NPQ adjusted for age and sex, fitted with limma with empirical Bayes moderation (trend and robust). The coefficient is the log2 difference (amnestic MCI minus non-amnestic MCI); fold change = 2^coefficient. FDR, Benjamini-Hochberg adjustment across all 127 proteins; p < 0.05 shown in bold. Proteins are ordered by p-value and use NULISA target names. These are the statistics plotted in the Figure 2 volcano plot. Abbreviations: FDR, false discovery rate; NPQ, NULISA Protein Quantification.

