## Supplementary figures and images for "Plasma Proteomic Identifies GDF15 as a Candidate Lithium Treatment Response Biomarker in Mild Cognitive Impairment: A Secondary Analysis of the LATTICE Randomized Clinical Trial"

### Supplemental Figure 1A

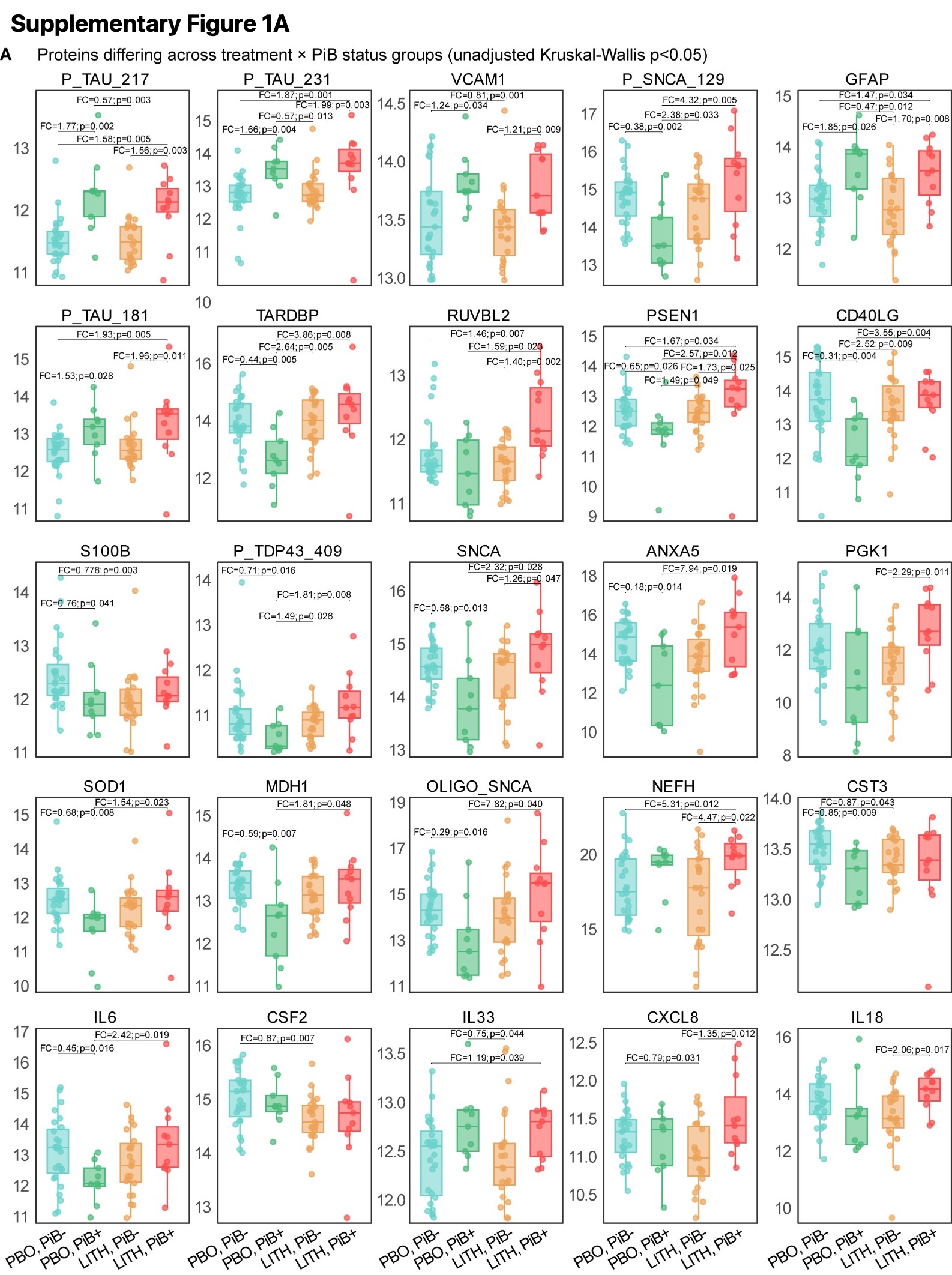

### Supplemental Figure 1B

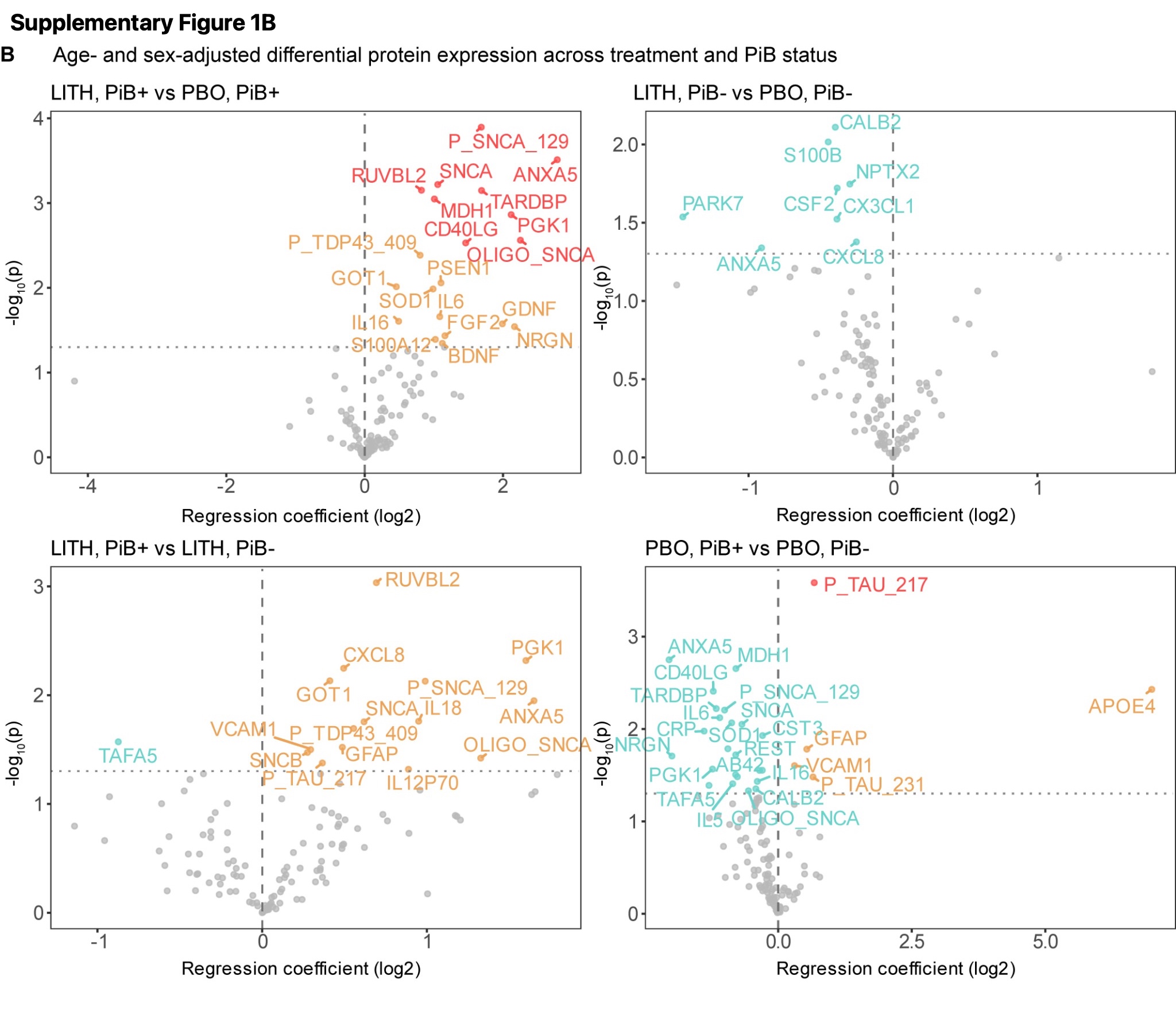

### Supplemental Figure 2

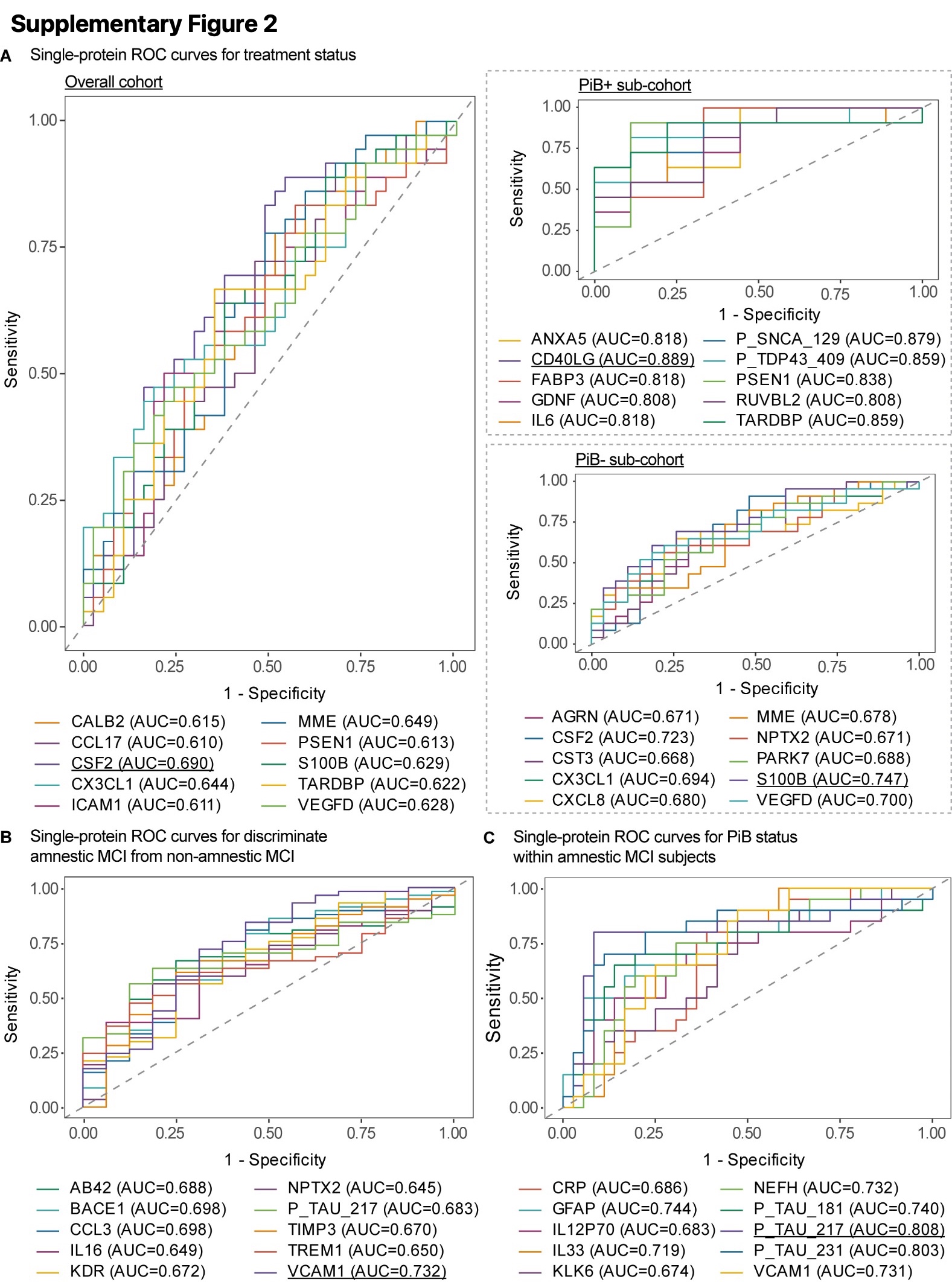

### Supplemental Figure 3

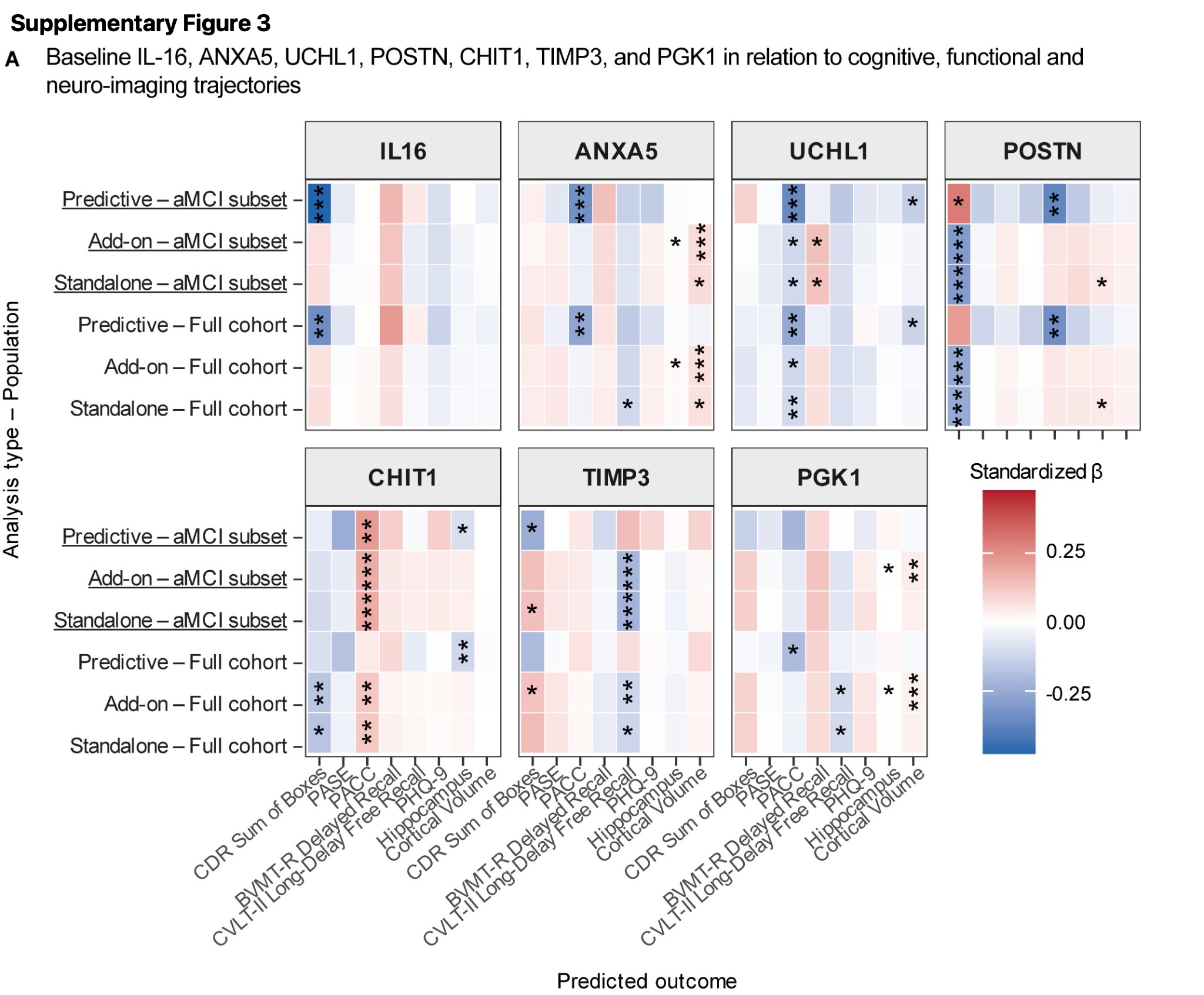
